# Genetic inactivation of zinc transporter SLC39A5 improves liver function and hyperglycemia in obesogenic settings

**DOI:** 10.1101/2021.12.08.21267440

**Authors:** Shek Man Chim, Kristen Howell, John Dronzek, Weizhen Wu, Cristopher Van Hout, Manuel Allen Revez Ferreira, Bin Ye, Alexander Li, Susannah Brydges, Vinayagam Arunachalam, Anthony Marcketta, Adam E Locke, Jonas Bovijn, Niek Verweij, Tanima De, Luca Lotta, Lyndon Mitnaul, Michelle G. LeBlanc, Regeneron Genetics Center, DiscovEHR collaboration, David Carey, Olle Melander, Alan Shuldiner, Katia Karalis, Aris N. Economides, Harikiran Nistala

## Abstract

Recent studies have revealed a role for zinc in insulin secretion and glucose homeostasis. Randomized placebo-controlled zinc supplementation trials have demonstrated improved glycemic traits in patients with type II diabetes (T2D). Moreover, rare loss-of-function variants in the zinc efflux transporter *SLC30A8* reduce T2D risk. Despite this accumulated evidence, mechanistic understanding of how zinc influences systemic glucose homeostasis and consequently T2D risk remains unclear. To further explore the relationship between zinc and metabolic traits, we searched the exome database of the Regeneron Genetics Center-Geisinger Health System DiscovEHR cohort for genes that regulate zinc levels and associate with changes in metabolic traits. We then explored our main finding using *in vitro* and *in vivo* models. We identified rare loss-of-function (LOF) variants (MAF<1%) in *Solute Carrier Family 39, Member 5 (SLC39A5)* associated with increased circulating zinc (p=4.9x10^-4^). Trans-ancestry meta-analysis across four studies exhibited nominal association of SLC39A5 LOF variants with decreased T2D risk. To explore the mechanisms underlying these associations, we generated mice lacking *Slc39a5*. *Slc39a5^-/-^* mice display improved liver function and reduced hyperglycemia when challenged with congenital or diet-induced obesity. These improvements result from elevated hepatic zinc levels and concomitant activation of hepatic AMPK and AKT signaling, in part due to zinc mediated inhibition of hepatic protein phosphatase activity. Furthermore, under conditions of diet-induced non-alcoholic steatohepatitis (NASH), *Slc39a5^-/-^* mice display significantly attenuated fibrosis and inflammation. Taken together, these results suggest SLC39A5 as a potential therapeutic target for non-alcoholic fatty liver disease (NAFLD) due to metabolic derangements including T2D.

**Lay summary:** Loss of the Zinc transporter SLC39A5 protects from obesity-driven hyperglycemia and liver pathology.

**Graphical abstract:** 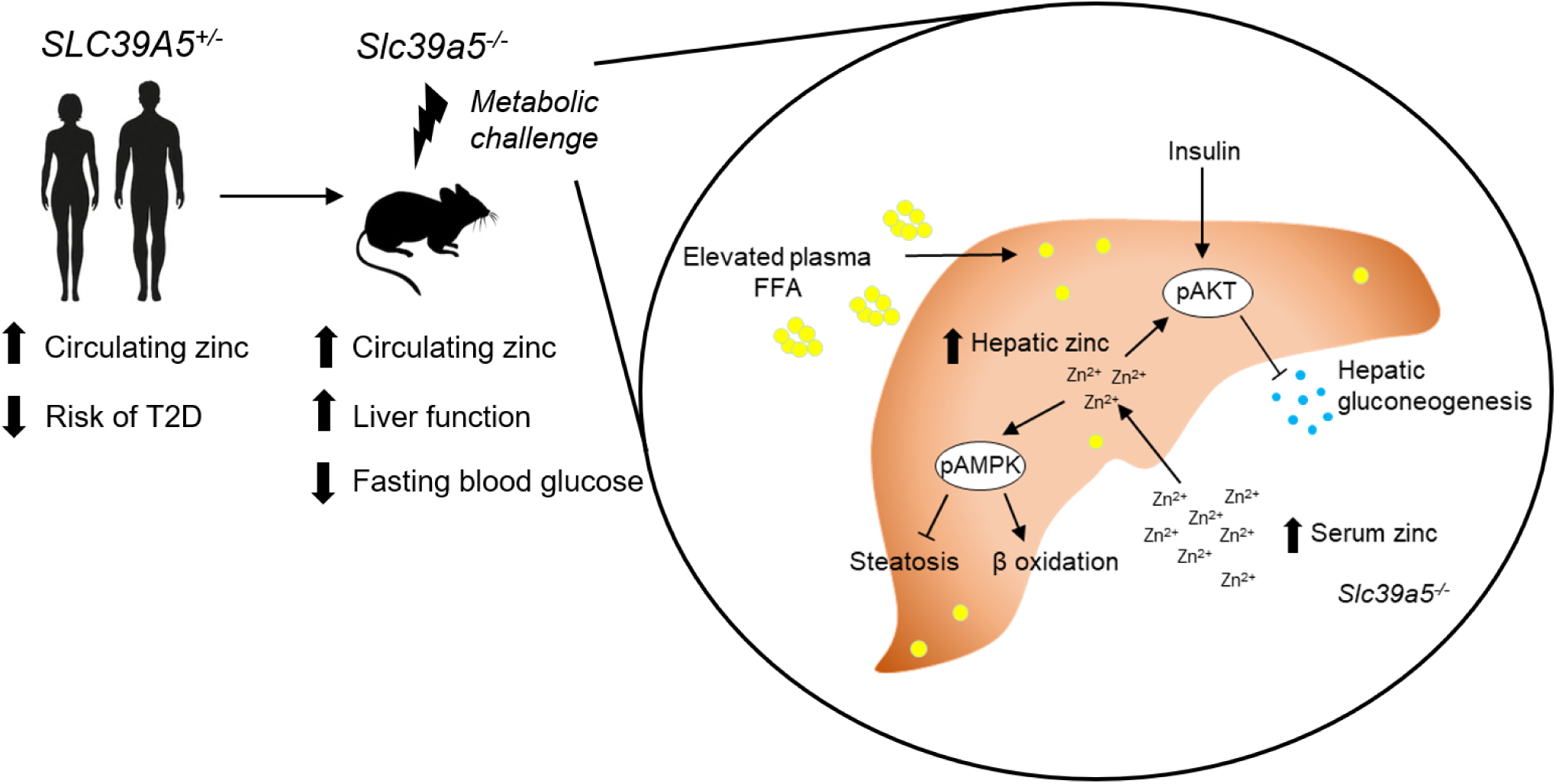

**Manuscript Highlights:**

- Heterozygous loss-of-function mutations in *SLC39A5* associated with elevated circulating zinc levels and nominal reduction in type II diabetes risk in humans.
- Loss of *Slc39a5* results in elevated circulating and hepatic zinc levels in mice.
- Mice lacking *Slc39a5* function are protected against hepatic steatosis and hyperglycemia resulting from diet-induced obesity or leptin-receptor deficiency and display reduced hepatic inflammation and fibrosis resulting from diet-induced NASH.
- Loss of *Slc39a5* function results in hepatic AMPK and AKT activation.
- SLC39A5 is a potential therapeutic target for fatty liver disease and type II diabetes.

## Introduction

Zinc (Zn^2+^) is an essential trace element with established roles in enzyme biochemistry and other biological processes. Hence, robust homeostatic mechanisms have evolved to maintain physiological levels of zinc and coordinate spatiotemporal demands across various tissues(*1*). Metal transporter proteins encoded by solute carrier (SLC) gene families SLC30 (zinc transporter, ZnT) and SLC39 (Zrt- and Irt-like protein, ZIP) facilitate zinc homeostasis by mediating cellular Zn^2+^ efflux and uptake respectively(*2*).

Converging lines of evidence have shown that zinc plays a crucial role in insulin secretion and glucose metabolism. For example, increasing zinc intake improves glycemic control in prediabetics and patients with T2D(*3*). Furthermore, loss-of-function (LOF) variation in *SLC30A8* (encoding ZnT8, a pancreatic islet zinc transporter) in humans associates with reduced glucose levels and a 65% reduction in T2D risk resulting from enhanced insulin responsiveness to glucose combined with increased pro-insulin processing(*4, 5*). To further explore mechanisms underlying the T2D-protective role of zinc and identify additional genetic determinants influencing systemic zinc homeostasis, we tested loss-of-function variation in zinc transporters for association with circulating zinc and T2D risk and identified rare putative LOF (pLOF) variants (MAF<1%) in *SLC39A5* associated with elevated circulating zinc (p=4.9x10^-4^). We demonstrate that the identified pLOF variants encode non-functional SLC39A5 proteins. In mice, loss of *Slc39a5* results in elevated hepatic zinc, lower glucose levels, and has protective effects in models of congenital and diet-induced obesity. These effects appear to be mediated by the activation of hepatic AMPK and AKT signaling, thereby uncovering a mechanistic basis for zinc induced liver protection and indicating that SLC39A5 inhibition may hold therapeutic potential in NAFLD and T2D.

## Results

### Rare loss-of-function variants in *SLC39A5* associate with elevated serum zinc and protection from type II diabetes

Using exome sequence data from participants of European ancestry in the Regeneron Genetics Center-Geisinger Health System DiscovEHR study, we identified rare pLOF variants (MAF<1%) in *SLC39A5* associated with increased circulating zinc levels in heterozygous carriers (p=4.9x10^-4^; Fig. 1A). We also tested rare LOF variants in *SLC39A5* for association with T2D in a multi-ethnic meta-analysis of four studies (UK Biobank, DiscovEHR, Mount Sinai’s BioMe study, and Malmö Diet and Cancer Study), totaling >62,000 cases and >518,000 controls, and found them to be nominally associated with protection from T2D (OR 0.82, 95%CI 0.68-0.99, p=3.7x10^-2^, Fig. 1B). Using serum call back analyses, we confirmed that circulating zinc levels in *SLC39A5* heterozygous loss of function carriers are elevated by 12% as compared to age, sex, BMI-matched reference controls (p=0.0024; Fig. 1C). Analyses of insulin production (proinsulin/insulin), insulin clearance (insulin/c-peptide ratio) and blood glucose demonstrated no differences based on genotype (Fig. 1D-I and Suppl. Table 1). These results, in conjunction with lack of *SLC39A5* expression in pancreatic β-cells(*6, 7*), suggest that SLC39A5 does not influence pancreatic β-cell development or function.

**Fig. 1.**
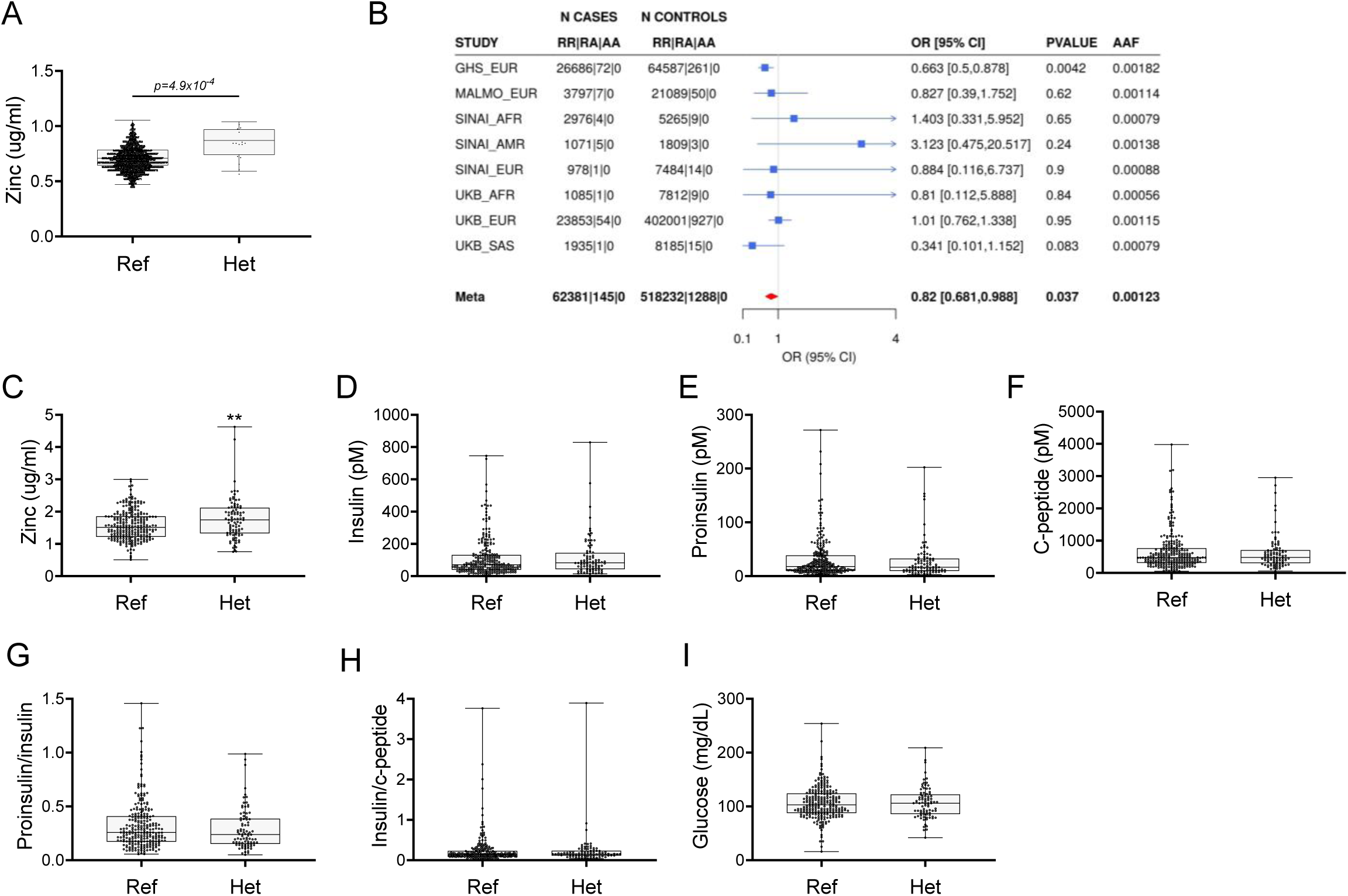
Rare pLOF variants in *SLC39A5* are associated with elevated serum zinc and nominal protection against type II diabetes (T2D). (A) Serum zinc in carriers of *SLC39A5* pLOF variants in the discovery cohort. Controls (Ref; *SLC39A5^+/+^*) and heterozygous carriers of pLOF variant alleles in *SLC39A*5 (Het; *SLC39A5^+/-^*). Subject numbers: Ref and Het respectively: n=5317 and n=15. (B) Trans-ancestry meta-analysis of association of *SLC39A5* pLOF variants with T2D. (C-I) Serum zinc and insulin profile of age, sex and BMI-matched controls in serum call back study. Subject numbers: Ref and Het respectively: n=246-253 and n=86-91, **P < 0.01, unpaired t-test. Numeric data is summarized in Suppl. Table 1.

To test whether the pLOF variants result in loss of protein function, we first examined their expression and cellular localization by immunofluorescence and flow cytometry. In these analyses we included several observed pLOF variants: p.Y47*(c.141C>G), p.R311*(c.931C>T), and p.R322*(c.964C>T). Bicistronic (IRES-DsRED) mammalian expression constructs encoding untagged wild-type or SLC39A5 muteins (Y47*, R311*, R322*,) were transfected into HEK293 cells (Fig. S1A-E). Consistent with previous reports(*8*), flow cytometry and immunofluorescence analyses at steady state demonstrated that wild-type SLC39A5 localized to the cell surface (Fig. S1A and B). In contrast, localization of variants Y47*, R311*, R322* to the cell surface was reduced by ∼91%, 98% and 99% respectively (Fig. S1D). To assess the zinc transport function of these variants, we leveraged a zinc-dependent transactivation assay using a metal regulatory element (MRE) responsive luciferase reporter. Wild-type SLC39A5 resulted in dose-dependent activation of the reporter to Zn^2+^ (an effect that was attenuated by zinc chelator N,N,N’,N’-Tetrakis(2-pyridylmethyl)ethylenediamine) (Fig. S1C), whereas variants Y47*, R311*, R322* failed to mediate a response (Fig. S1D and E). Therefore, variants Y47*, R311*, R322* encode non-functional proteins. Their association with elevated serum zinc levels in the corresponding carriers is consistent with the proposed role of SLC39A5 in maintaining systemic zinc homeostasis by facilitating efflux of excess serosal zinc into the gut lumen(*8*).

### *Slc39a5* homozygous-null mice display elevated serum and tissue zinc

To investigate the role of *Slc39a5* in glucose homeostasis *in vivo*, we generated *Slc39a5*-null mice (Fig. S2A). The resulting *Slc39a5^-/-^* mice completely lacked *Slc39a5* transcript and protein in their duodenum and liver (Fig. S2B and C), two tissues with documented expression of SLC39A5(*8*). Consistent with our observations in human heterozygous LOF carriers, *Slc39a5^+/-^* mice had elevated circulating zinc levels (∼26% in female and ∼23% in male) compared to wildtype littermates. The elevation in circulating zinc was greatly accentuated in *Slc39a5^-/-^* mice (∼280% in females and ∼227% in males) compared to wild-type littermates (Fig. 2A). *Slc39a5^-/-^* mice displayed normal fecundity and had no overt phenotypes even at 22 months of age. Elemental analyses of major organs (in both sexes) revealed that *Slc39a5^-/-^* mice had significantly elevated zinc levels in the liver, bone, kidneys and brain, and lower levels in the pancreas (Fig. 2B and Suppl. Table 2). These phenotypes are consistent with previously reported *Slc39a5* knockout mouse models(*9, 10*). No differences in magnesium, iron, copper, cobalt or calcium were observed in liver (Fig. S11C and F). Serum chemistry analysis in adult mice (10 months, both sexes) demonstrated no differences in pancreatic amylase, renal function parameters (blood urea nitrogen, creatinine, total protein and uric acid), electrolytes (chloride, potassium and sodium), and liver enzymes (alanine aminotransferase; ALT and aspartate aminotransferase; AST) (Suppl. Table 3), suggesting that the observed changes in tissue zinc levels are physiologically inert at this age. Unlike *Slc39a5^-/-^* mice, *Slc39a5^+/-^* mice showed no changes in tissue zinc levels despite elevation in serum zinc indicating that the free exchangeable pool of serum zinc in the *Slc39a5^+/-^* mice is not sufficient to alter zinc balance within tissues (Suppl. Table 2). Conservation at the protein level (∼83.5% identity), similar postnatal expression(*11*) and preserved function between mouse and human orthologs suggest that *Slc39a5^-/-^* mice provide a valid model to explore the observed subthreshold T2D protective effect of *SLC39A5* LOF alleles in humans.

**Fig. 2.**
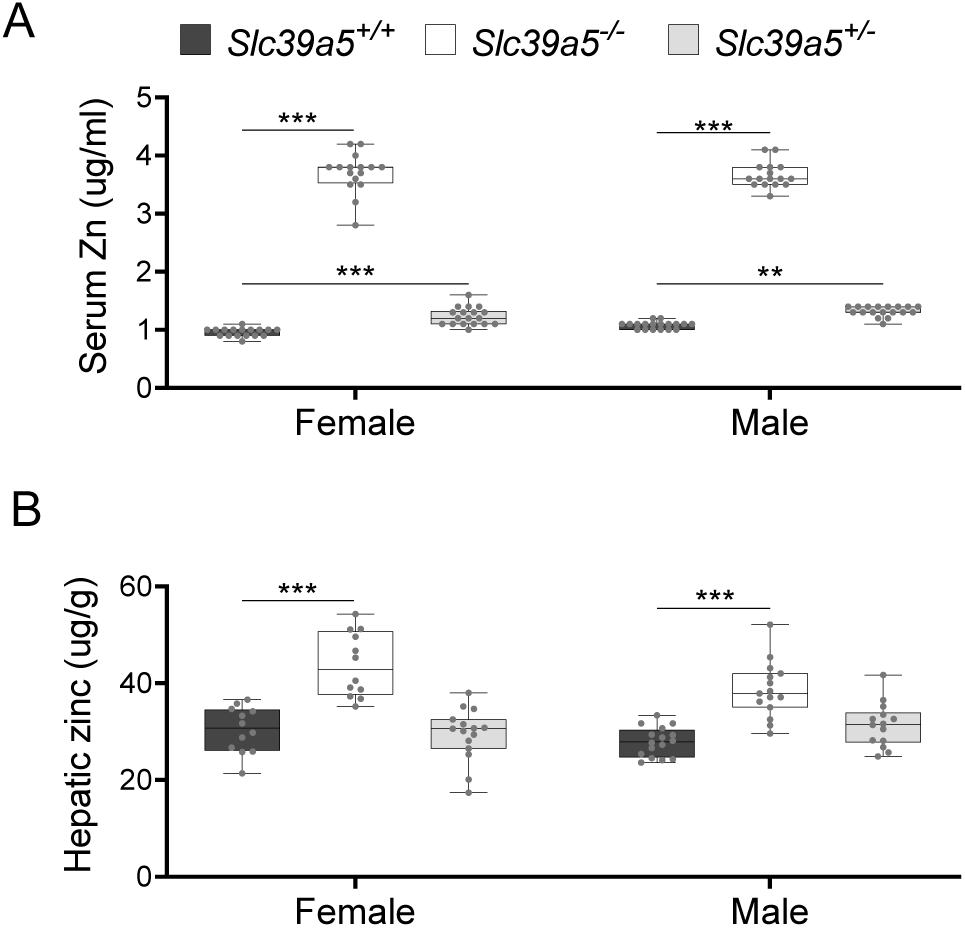
Loss of *Slc39a5* results in elevated circulating and hepatic zinc levels in mice. Serum zinc (A) and hepatic zinc (B) in *Slc39a5*^+/+^*, Slc39a5*^-/-^ and *Slc39a5*^+/-^ mice at 40 weeks of age, n=16-18. **P < 0.01, ***P < 0.001, two-way ANOVA with post hoc Tukey’s test.

### Loss of *Slc39a5* results in reduced fasting blood glucose in congenital and diet-induced obesity models

To assess whether disruption of Slc39a5 function improves glycemic traits in mice, we challenged the *Slc39a5^-/-^* mice with well-established models of congenital (leptin-receptor deficiency; *Lepr^-/-^* mice) or diet induced obesity(*12, 13*). Loss of *Slc39a5* did not alter body weight in either model (Fig. 3A, E, I, M and Fig. S4A, S5A, S6A, S7A). *Slc39a5^-/-^*;*Lepr^-/-^* mice or *Slc39a5^-/-^* mice on high fat high fructose diet (HFFD) showed significant reduction in fasting blood glucose levels as compared to littermate controls (Fig. 3B, F, J, N and Suppl. Table 4 and 5), but not fasting insulin levels (Fig. 3C, G, K, O). However, *Slc39a5^+/-^* mice did not show a similar improvement in fasting blood glucose (Fig. S2D and E), indicating that loss of one copy of *Slc39a5* does not actuate a protective glucose lowering mechanism; hence, we leveraged *Slc39a5^-/-^* mice for further mechanistic exploration.

**Fig. 3.**
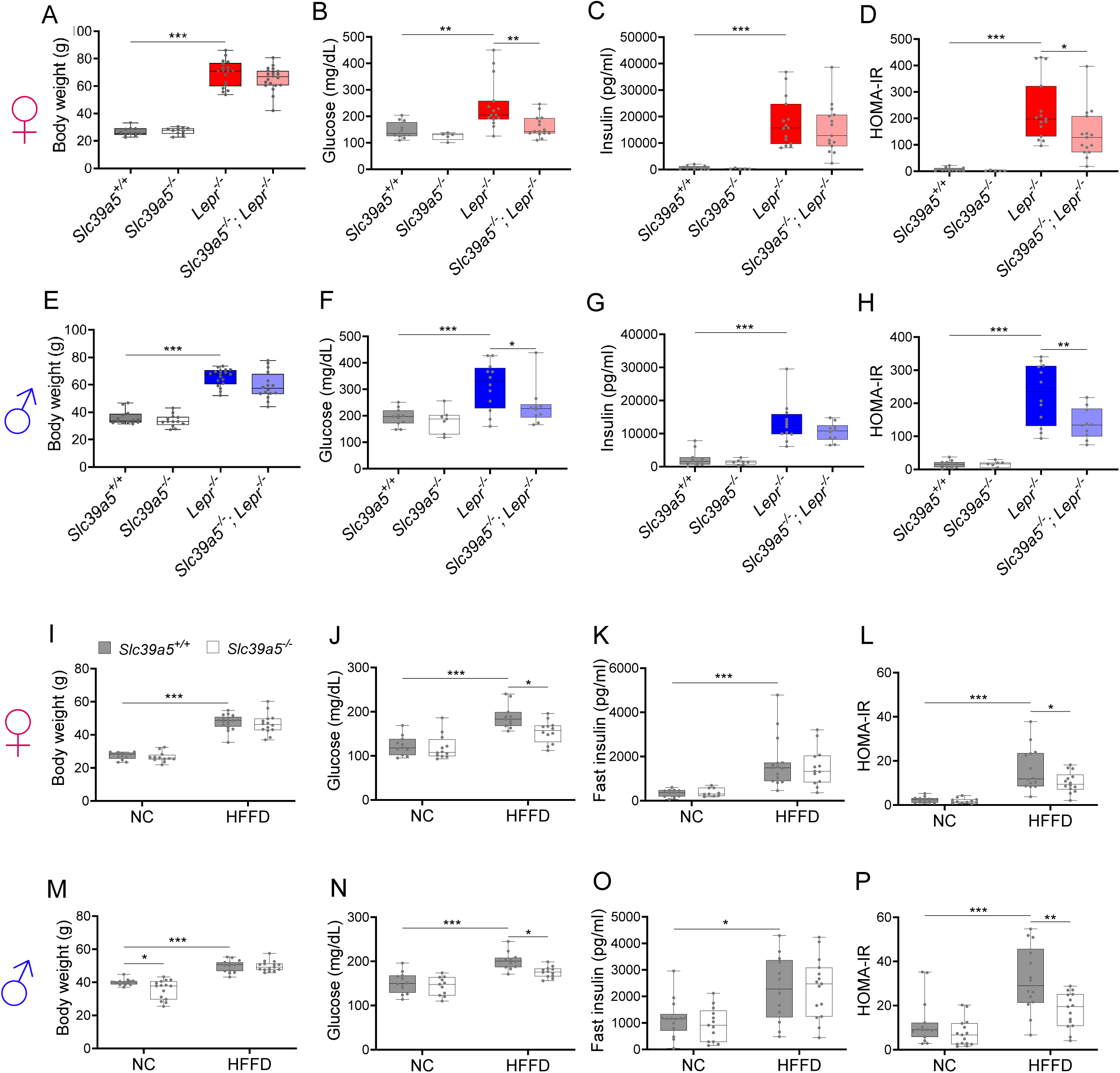
Loss of *Slc39a5* improves glycemic traits in leptin-receptor deficient mice and in mice challenged with high fat high fructose diet (HFFD). Female (A-D, I-L; ♀) and Male (E-H, M-P; ♂) mice. (A-H) *Slc39a5^-/-^;Lepr^-/-^* and corresponding control mice. (A, E) Body weight at 34 weeks. (B, F) Fasting blood glucose at 34 weeks. (C, G) Fasting insulin at 34 weeks. (D, H) Homeostatic model assessment for insulin resistance (HOMA-IR) at 34 weeks. *Slc39a5*^+/+^ *and Slc39a5*^-/-^ (n=5-12), *Lepr* ^-/-^ and *Slc39a5* ^-/-^;*Lepr* ^-/-^ (n=10-15). *P < 0.05, **P < 0.01, ***P < 0.001, one-way ANOVA with post hoc Tukey’s test. (I-P) *Slc39a5^-/-^ and Slc39a5^+/+^* mice were fed HFFD or NC for 30 weeks. (I, M) Body weight at 30 weeks. (J, N) Fasting blood glucose at 30 weeks. (K, O) Fasting insulin at 30 weeks. (L, P) HOMA-IR at 30 weeks, n=11-15. *P < 0.05, **P < 0.01, ***P < 0.001, two-way ANOVA with post hoc Tukey’s test. Numeric data is summarized in Suppl. Table 4 and 5.

Loss of *Slc39a5* in these models demonstrated improved glucose tolerance despite no differences in insulin secretion or clearance at steady state (except female *Slc39a5^-/-^; Lepr^-/-^* upon fasting) as compared to littermate controls (Fig. S3, S4E-F, S5E-F, S6E-F, S7E-F, S8A-H). Consistently, loss of *Slc39a5* resulted in reduced insulin resistance in these models (Fig. 3D, H, L, P). Consistent with these observations, no differences in insulin production or clearance were observed in heterozygous carriers of *SLC39A5* LOF variants as compared to age, sex, BMI-matched reference controls (Fig. 1D-H) upon serum call back analyses. Combined with the fact that single-cell transcriptomic data in both human and mouse show no expression of *SLC39A5* in pancreatic β-cells (*6, 7, 14, 15*), these results indicate that the glucose lowering effects in *Slc39a5^-/-^* mice appear to be independent of pancreatic β-cell function.

### Loss of *Slc39a5* improves liver function

Given that NAFLD and T2D are concurrent comorbidities characterized by hepatic steatosis, glucose intolerance and insulin resistance(*16*), we explored whether loss of *Slc39a5* and consequent hepatic zinc accumulation (Fig. 2B) influenced liver function in models of congenital obesity and diet-induced obesity.

*Slc39a5^-/-^;Lepr^-/-^* mice displayed significant reductions in hepatic lipid accumulation (Fig. 4A and Fig. S9A), hepatic triglyceride content (Fig. 4B and Fig. S9B), and in serum ALT and AST levels (biomarkers of liver damage) (Fig. 4C, D, Fig. S9C, D and Suppl. Table 4) compared to littermate *Lepr^-/-^* mice. Moreover, *Slc39a5^-/-^; Lepr^-/-^* mice displayed reduced NAFLD activity score (an aggregate score of macrovesicular steatosis, hepatocellular hypertrophy and inflammation) (Fig. 4E and Fig. S9E). Consistent with reduced lipid burden, expression of hepatic fatty acid synthase expression trended lower in *Slc39a5^-/-^*;*Lepr^-/-^* mice (Fig. S4B-D, S5B-D). Moreover, hepatic and serum beta-hydroxybutyrate levels were elevated in *Slc39a5^-/-^*; *Lepr^-/-^* mice as compared to *Lepr^-/-^* mice, indicative of elevated mitochondrial β-oxidation and disposal of excess hepatic lipid resulting from the leptin receptor deficiency (Fig. 4F, Fig. S8K-L, S9F).

**Fig. 4.**
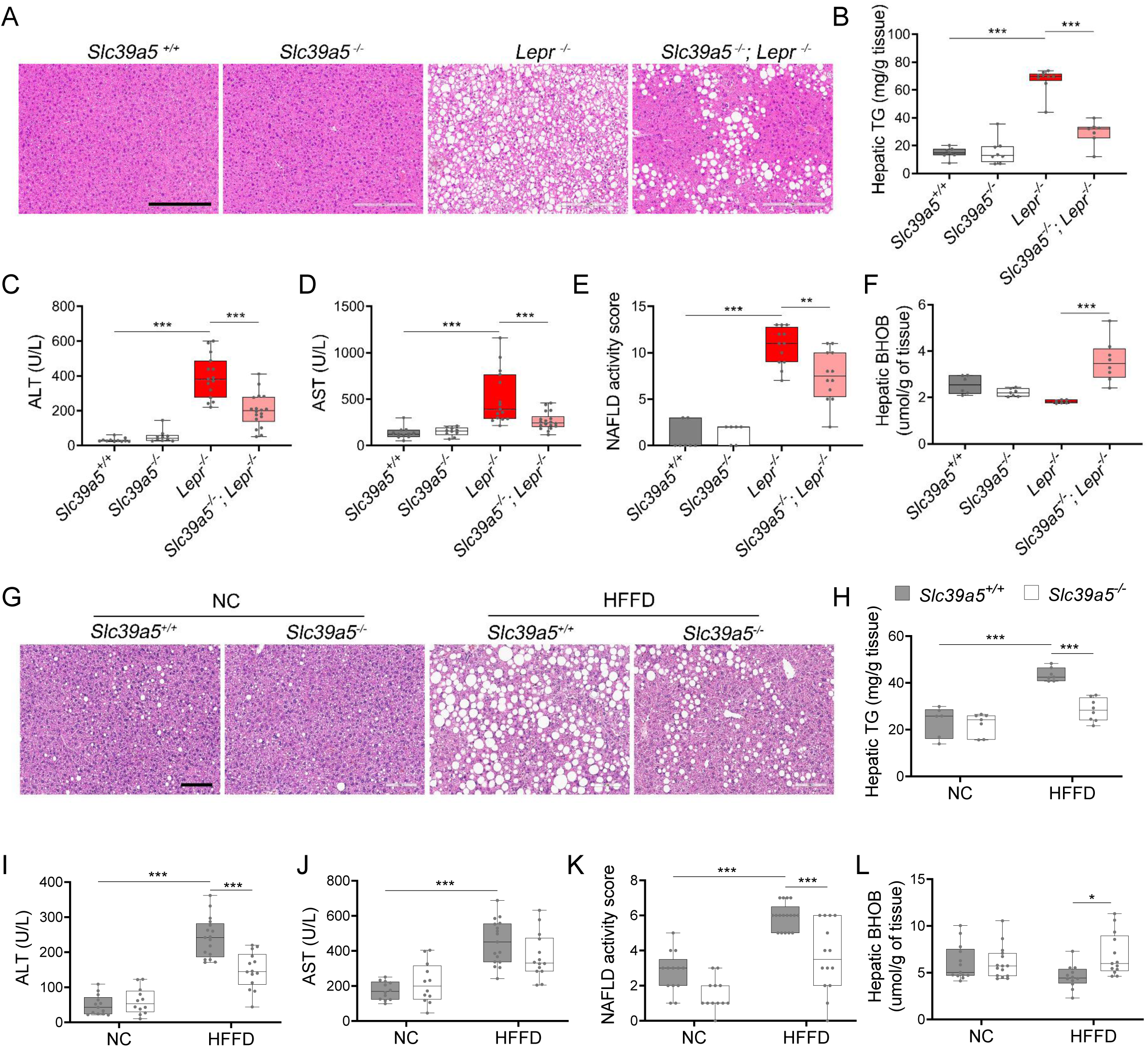
Loss of *Slc39a5* improves liver function and steatosis in leptin-receptor deficient female mice and in female mice challenged with high fat high fructose diet (HFFD). *Slc39a5^-/-^;Lepr^-/-^* and corresponding control mice (A-F) were sacrificed after 16 hour fasting at 34 weeks of age. (G-L) *Slc39a5^-/-^ and Slc39a5^+/+^* mice were fed HFFD or NC for 30 weeks and sacrificed after 16 hours fasting. (A, G) Representative images of livers stained with H&E. Scale bar, 200µm. (B, H) Hepatic triglyceride (TG) content in explanted liver samples at endpoint. (C, I) Serum ALT. (D, J) Serum AST. (E, K) NAFLD activity score, (F, L) Hepatic beta-hydroxybutyrate (BHOB). *P < 0.05, **P < 0.01, ***P < 0.001, *Slc39a5^-/-^;Lepr^-/-^*and corresponding control mice: one-way ANOVA with post hoc Tukey’s test, HFFD or NC: two-way ANOVA with post hoc Tukey’s test. Numeric data is summarized in Suppl. Table 4 and 5.

Next, we examined whether loss of *Slc39a5* improves liver function in diet-induced obesity. HFFD significantly increased body weight, serum ALT and AST levels, and NAFLD activity score (Suppl. Table 5). Loss of *Slc39a5* had no considerable impact on body weight in this model but resulted in marked reductions of hepatic triglyceride content in both sexes (Fig. 4H and S9H). However, loss of *Slc39a5* resulted in sex-specific differences in most NAFLD related traits, with females benefiting more significantly compared to males, displaying significant reductions in hepatic steatosis (Fig. 4G), serum ALT (but not AST) (Fig. 4I and J), NAFLD activity score (Fig. 4K), and hepatic fatty acid synthase levels (Fig. S6B-D), and a significant elevation in hepatic and serum beta-hydroxybutyrate levels (Fig. 4L and Fig. S8J). Lastly, in contrast to what was observed in *Slc39a5^-/-^*;*Lepr^-/-^* mice, hepatic glucose-6-phosphatase levels were significantly reduced in HFFD female *Slc39a5^-/-^* mice (Fig. S6B-D), suggesting that reduced hepatic gluconeogenesis may contribute in part to the observed glucose lowering in these mice.

In HFFD male *Slc39a5^-/-^* mice, however, there were no improvements in serum ALT, AST and NAFLD activity score (Fig. S9I-K), despite reductions in hepatic triglyceride content (Fig. S9H). Significant elevation in hepatic and serum beta-hydroxybutyrate levels and nominal reduction in hepatic fatty acid synthase levels (Fig. S7B-D, S8J and S9L) were suggestive of reduced lipid burden in HFFD male *Slc39a5^-/-^* mice.

Taken together, these studies suggest that loss of *Slc39a5* in metabolically challenged mice results in reduced hepatic lipid burden and improved hepatic insulin sensitivity, ultimately leading to improved systemic glucose homeostasis.

### Loss of *Slc39a5* results in activation of hepatic AMPK and AKT signaling

To explore the mechanism underlying improved hepatic steatosis and glycemic traits in *Slc39a5^-/-^* mice we evaluated two key signaling hubs that mediate lipid metabolism and insulin sensitivity, AMPK and AKT. Activation of hepatic AMPK signaling in a diet-induced obesity model reduces hepatic steatosis and downstream inflammation and fibrosis(*17*), whereas activation of hepatic AKT reduces glucose production in liver(*18*). Hence, we evaluated phosphorylation of Thr172 in AMPKα subunit (p.AMPKα) and Ser473 phosphorylation of AKT (p.AKT) in liver lysates from *Slc39a5^-/-^*; *Lepr^-/-^* mice and HFFD-fed *Slc39a5^-/-^* mice and their respective controls. Prior to evaluating p.AMPKα and p.AKT, we confirmed that all *Slc39a5^-/-^* mice used in these experiments displayed hepatic zinc accumulation (Fig. 5B and E, Fig. S10B and E) and increased expression of the zinc-responsive genes *Mt1* and *Mt2* (Fig. 5C and F, Fig. S10C and F, S11B and E, S12B and E). The p.AMPKα levels were elevated in both *Slc39a5^-/-^*; *Lepr^-/-^* (Fig. 5A and Fig. S10A and 11A and D) and HFFD *Slc39a5^-/-^* mice (Fig. 5D and Fig. S10D and S12A and D) regardless of sex as compared to controls. However, significant hepatic AKT activation was observed only in female *Slc39a5^-/-^*; *Lepr^-/-^* and HFFD *Slc39a5^-/-^* mice (Fig. 5A and D, Fig. S10A and D, S11A and D, S12A and D).

**Fig. 5.**
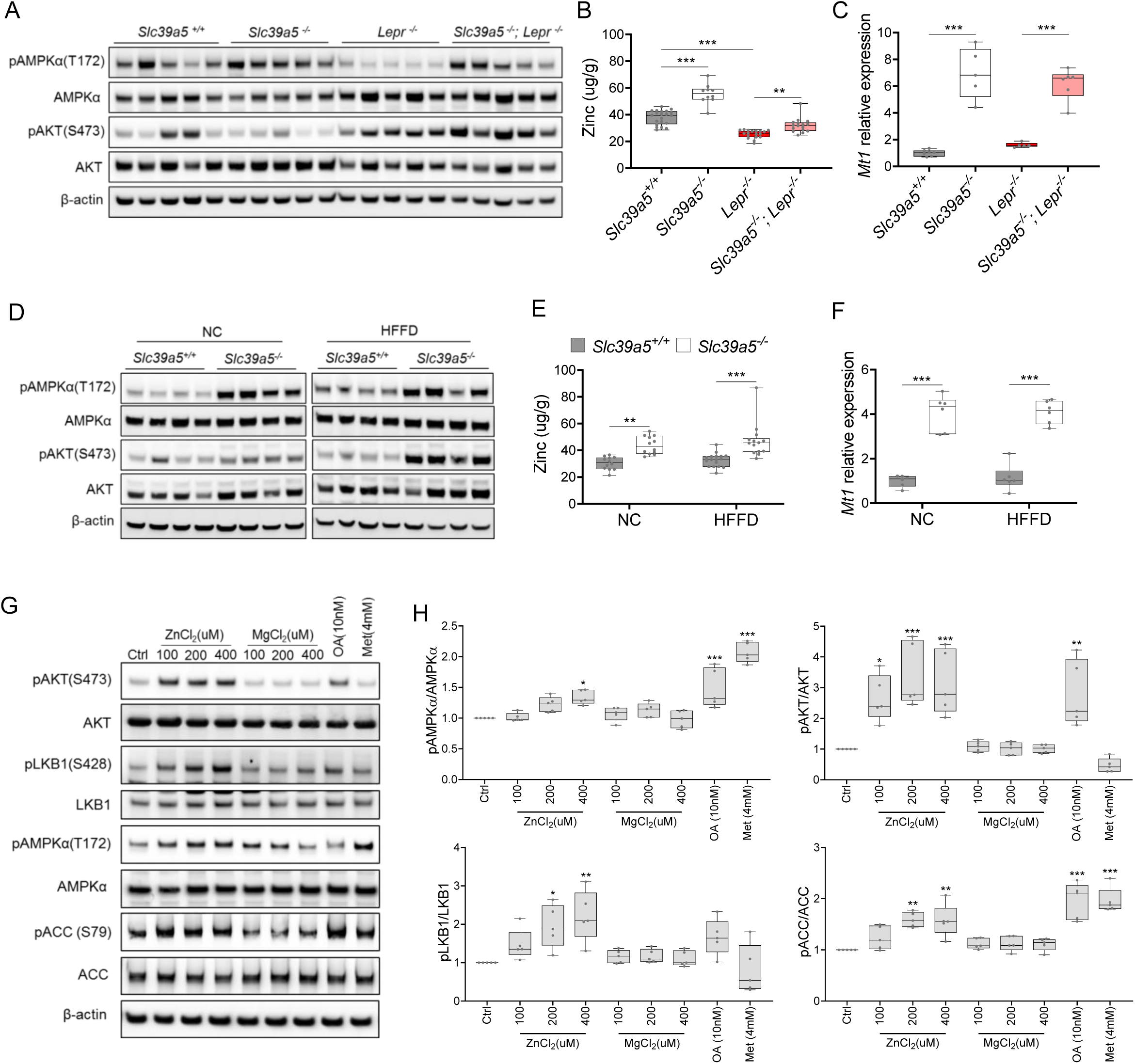
Loss of *Slc39a5* results in elevated hepatic zinc and activation of hepatic AMPK signaling in leptin-receptor deficient female mice and female mice challenged with high fat high fructose diet (HFFD). Analyses were done on explanted liver samples collected after 16 hours fasting at endpoint in *Lepr^-/-^* (A-C) and HFFD mice (D-F). (A, D) Immunoblot analysis of hepatic AMPK and AKT activation. AMPK and AKT signaling is activated in *Lepr^-/-^*;*Slc39a5^-/-^* mice and HFFD *Slc39a5^-/-^* mice (compared to their *Scl39a5^+/+^* counterparts). (B, E) Hepatic zinc is elevated in *Lepr^-/-^*;*Slc39a5^-/-^* mice and HFFD *Slc39a5^-/-^* mice (n=10-21). (C, F) Elevated hepatic zinc results in increased *Mt1* (zinc responsive gene) expression in both models. (G) Immunoblot analysis of primary human hepatocytes treated with zinc chloride (ZnCl_2_), magnesium chloride (MgCl_2_), okadaic acid (OA), metformin (Met) for 4 hours. Zinc activated AMPK and AKT signaling in primary human hepatocytes. (H) Densitometric analysis of immunoblots (compared to control). *P < 0.05, **P < 0.01, ***P < 0.001, ANOVA with post hoc Tukey’s test.

To further explore the potential role of elevated hepatic zinc in AMPK and AKT activation, we examined whether exogenous zinc activates AMPK and AKT signaling in primary human hepatocytes. Zinc activated AKT signaling in these cells in a dose dependent manner with no adverse effect on cell viability, whereas magnesium had no effect (Fig. 5G and H, Fig. S13A). Moreover, zinc activated AMPK signaling and its downstream substrates acetyl-CoA carboxylase (ACC), and liver kinase B1 (LKB1; the kinase responsible for AMPKα Thr172 phosphorylation) (Fig. 5G and H). Time-resolved analyses of zinc-mediated activation of LKB, AMPK, and AKT indicated that zinc activates AMPK and AKT signaling acutely (within 4 hours) suggesting that zinc influences phosphorylation of these substrates independent of *de novo* protein synthesis (Fig. S13B). Similar results were obtained in the human hepatoma cell line HepG2 (Fig. S13C and D).

Zinc is a potent inhibitor of protein phosphatases, including protein phosphatase 2A (PP2A) and protein tyrosine phosphatase-1B (PTP1B)(*19, 20*), both of which regulate the phosphorylation of AMPKα. Liver-specific ablation of *Ppp2cα* (encoding PP2A’s catalytic subunit) improves glucose tolerance and insulin sensitivity in mice(*21*), whereas liver-specific ablation of *Ptpn1* (encoding PTP1B) improves glucose tolerance, insulin sensitivity, and lipid metabolism(*22*). Given that hepatic zinc is elevated in *Slc39a5^-/-^* mice, we evaluated hepatic phosphoserine/threonine (p.Ser/Thr) and phosphotyrosine (p.Tyr) phosphatase activity in the congenital and diet-induced obesity mice at endpoint. *Slc39a5^-/-^*;*Lepr^-/-^* mice displayed reduced p.Ser/Thr and p.Tyr phosphatase activity compared to *Lepr^-/-^* littermate controls (Fig. S14A and B). Under HFFD, female *Slc39a5^-/-^* mice showed reduced hepatic p.Ser/Thr and p.Tyr phosphatase activity (33% and 28% respectively), and non-statistically significant reductions were also observed in male *Slc39a5^-/-^* mice (Fig. S14C and D). Consistent with these observations, exogenous zinc inhibited p.Ser/Thr and p.Tyr phosphatase activity in primary human hepatocytes in a dose dependent manner (Fig. S14E). These results point to zinc mediated inhibition of protein phosphatase activity as a likely mechanism underlying hepatic AMPK and AKT activation in *Slc39a5^-/-^* mice.

### Loss of *Slc39a5* reduces hepatic inflammation and fibrosis upon a NASH dietary challenge

NAFLD encompasses a continuum of liver conditions from nonalcoholic fatty liver characterized by steatosis, to nonalcoholic steatohepatitis (NASH) characterized by inflammation and fibrosis(*23*). The improvements in liver function and steatosis in congenital and diet-induced obesity mouse models lacking *Slc39a5*, led us to investigate whether loss of *Slc39a5* protects against NASH. Diet-induced NASH significantly increased serum ALT and AST levels (Fig. 6A and B, Fig. S15A and B), body weight, fasting blood glucose (Fig. S16A, B, F, G) and liver fibrosis (Fig. 6H and I, Fig. S15H and I) in *Slc39a5^+/+^* mice (Suppl. Table 6). In contrast, *Slc39a5^-/-^* mice challenged with diet-induced NASH displayed significant reductions in serum ALT and AST levels (Fig. 6A and B, Fig. S15A and B) and fasting blood glucose (Fig. S16B and G), along with significant improvements in hepatic inflammation and fibrosis (Fig. 6E and H, Fig. S15E and H) and the expected increases in serum and hepatic zinc (Fig. S16C, D, H, I). Consistently, hepatic collagen deposition (Fig. 6G, Fig. S15G) were significantly reduced in NASH *Slc39a5^-/-^* mice. However, NASH *Slc39a5^-/-^* mice were not protected from hepatic steatosis or hepatocyte hypertrophy (Fig. 6C and D, S15C and D). NAFLD activity score and steatosis-activity-fibrosis score (sum of NAFLD activity score and fibrosis score) were significantly reduced in female NASH *Slc39a5^-/-^* mice, but not in their male counterparts (Fig. 6F and I, Fig. S15F and I). Nonetheless, hepatic superoxide dismutase (SOD) activity was significantly elevated in both sexes in NASH *Slc39a5^-/-^* mice (Fig. S16E and J), suggesting that the increase in hepatic zinc may be ameliorating the increased hepatic oxidative stress observed in NASH(*23*).

**Fig. 6.**
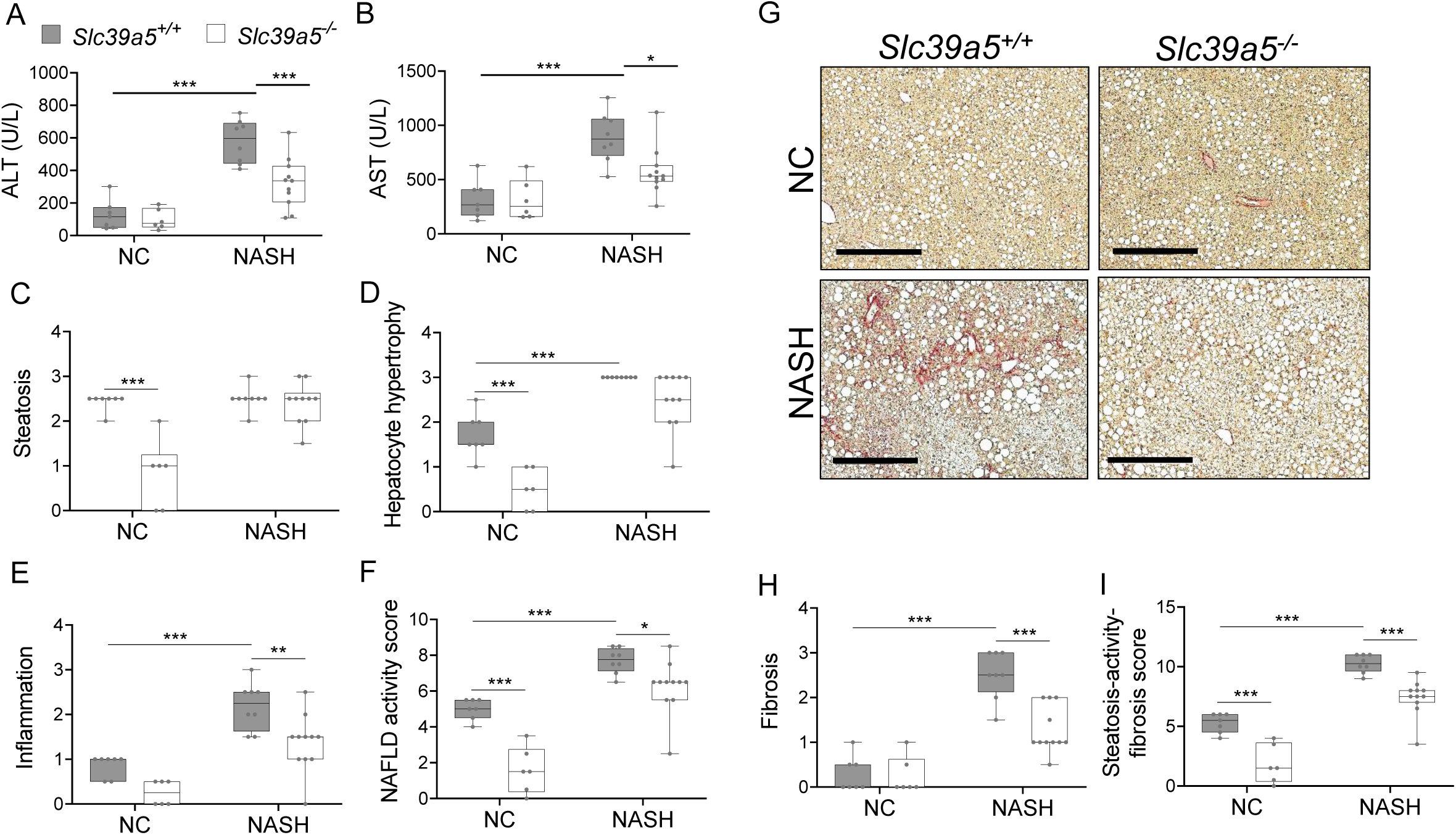
Loss of *Slc39a5* improves hepatic inflammation and fibrosis in female mice challenged with diet-induced NASH. *Slc39a5^-/-^ and Slc39a5^+/+^* mice were placed on a NASH inducing diet or NC for 40 weeks and sacrificed after 16 hours of fasting. (A, B) NASH *Slc39a5^-/-^* mice display reduced serum ALT and AST levels. (C-E) Histology scores for steatosis, hepatocyte hypertrophy, inflammation. (F) NAFLD activity score was reduced in NASH *Slc39a5^-/-^* mice. (G-I) NASH *Slc39a5^-/-^* mice display reduced fibrosis. (G) Representative images of explanted livers sample stained with picrosirius red indicative of collagen deposition. Scale bar, 300µm. (H, I) Fibrosis and steatosis-activity-fibrosis scores. n=6-7 (NC) and 8-11 (NASH), *P < 0.05, **P < 0.01, ***P < 0.001, two-way ANOVA with post hoc Tukey’s test. Numeric data is summarized in Suppl. Table 6.

In aggregate, these studies suggest that the favorable metabolic profile in the *Slc39a5^-/-^* mice results from convergent hepatoprotective effects due to reduced lipotoxic and oxidative stress.

## Discussion

Zinc is a required trace element for many biological processes. Hence, homeostatic mechanisms have evolved to maintain optimal zinc levels across tissues(*1*). This regulation is accomplished by multiple transporters encoded by the *SLC30* and *SLC39* gene families(*2*). Given the apparent complexity of the system, we chose to take a human genetics approach to search for zinc transporter genes associated with metabolic traits and discovered a novel association of LOF variants in *SLC39A5* with increased circulating zinc (p=4.9x10^-4^) and a reduced risk of T2D (OR 0.82, 95%CI 0.68-0.99, p=3.7x10^-2^).

To firm up this association and explore underlying molecular mechanisms, we generated mice lacking *Slc39a5*. In line with the human data, and consistent with the proposed role of *Slc39a5* as a non-redundant cell surface zinc transporter facilitating endogenous zinc excretion(*8, 9, 11*), there was significant zinc accumulation in *Slc39a5^-/-^* mice across several tissues including liver (Suppl. Table 2). However, there was no significant accumulation of zinc in tissues of *Slc39a5* heterozygous-null mice on a zinc adequate diet despite significant increases in serum zinc (∼26% in females and ∼23% in males; Fig. 2), suggesting that the relative increase of serum zinc in heterozygotes (albeit significant) is insufficient to increase zinc levels in tissues with substantial zinc stores such as the liver.

Nonetheless, given the connection with protective effects arising from heterozygous loss of *SLC39A5* in humans, we examined the effect of loss of *Slc39a5* in mice under conditions of metabolic stress, employing models of congenital or diet-induced obesity, and NASH. We demonstrate that in all three models, loss of *Slc39a5* (in homozygosis) has protective effects that arise from elevation of circulating and hepatic zinc levels. In congenital or HFFD-induced obesity, there was improvement in glycemic traits and liver function, and a reduction of steatosis, which were not accompanied by reductions in body weight or changes in insulin profile. In a model of diet-induced NASH, loss of *Slc39a5* reduced hepatic inflammation and fibrosis, but without significant changes in steatosis. Mechanistically, these protective effects result at least in part from inhibition of protein phosphatases (as result of elevated levels of zinc), and consequent increase in hepatic AMPK and AKT activation.

The observed protective metabolic effects appear to be extra-pancreatic in both mice and humans, as supported by several lines of evidence. Carriers of heterozygous LOF mutations in *SLC39A5* have elevated serum zinc but exhibit no differences in insulin production or clearance as compared to age, sex, and BMI-matched homozygous reference controls (Fig. 1C-E). As in humans, loss of *Slc39a5* in mice results in elevated serum zinc (Fig. 2A) without impairment in pancreatic function (Suppl. Table 3). Moreover, the observed antihyperglycemic effects in *Slc39a5^-/-^* mice are not driven by changes in insulin production or clearance (Fig. S4-S7). Taken together, these observations suggest that the protective metabolic changes are extra-pancreatic.

Furthermore, our data strongly indicates that the protective effects of loss of *Slc39a5* are actuated by elevated hepatic zinc concentrations. Several lines of evidence support this interpretation. First, metabolic challenges in the form of congenital or diet-induced obesity in mice revealed hepatic zinc deficiency (Fig. 5B, Fig. S10B) along with associated comorbidities including hepatic steatosis, increased fasting blood glucose and impaired insulin sensitivity (Fig. 3). Second, loss of *Slc39a5* in these models resulted in the accumulation of serum zinc and hepatic zinc and concomitant improvement in liver function (Fig. 4-5, Fig. S9) and systemic glucose homeostasis (Fig. 3, Fig. S3). This data are consistent with observations that zinc deficiency is associated with obesity(*24*) and is a biochemical hallmark of fatty liver disease in both rodents and humans(*25*); conversely, zinc supplementation reverses manifestations of zinc deficiency in fatty liver disease and long-term oral zinc supplementation can support liver function and prevent hepatocellular carcinoma development in patients with chronic liver diseases(*26*).

The importance of hepatic zinc in the protective effects against obesity and NASH is further supported by our findings that elevated hepatic zinc in *Slc39a5^-/-^* mice enhanced hepatic AMPK and hepatic AKT signaling (Fig. 5 and Fig. S10). These increases correlated with reductions in hepatic p.Ser/Thr phosphatase and p.Tyr phosphatase levels in both diet-induced and congenital obesity models (Suppl. Fig. S14A-D), and were corroborated by *in vitro* evidence (Fig. S14E-F). These findings mirror prior studies showing that zinc inhibits protein serine/threonine and tyrosine phosphatases that dephosphorylate AMPK and AKT (*19, 27–31*). In turn, in states of lipotoxic stress, serine/threonine phosphatases such as PP2A and PP2C inhibit AMPK resulting in a feed forward effect of the lipid overload(*32, 33*), whereas protein tyrosine phosphatases including PTP1B, TCPTP and PTEN have been implicated in systemic glucose homeostasis by regulating the PI3K-AKT pathway (*22, 30, 34, 35*).

Overall, our studies indicate that the favorable metabolic profile observed in the *Slc39a5^-/-^* mice results from the loss of endogenous zinc excretion and concomitant systemic zinc redistribution. Our study provides for the first-time genetic evidence demonstrating the protective role of zinc against hyperglycemia and unravels the mechanistic basis underlying this effect. Taken together, these observations suggest SLC39A5 inhibition as a potential therapeutic avenue for T2D, and other indications where zinc supplementation alone is inadequate.

## Materials and methods

*Slc39a5*^-/-^ and *Lepr^-/-^* mouse lines were created using VelociGene^®^ technology(*39*). *Lepr*^-/-^ mice were bred with *Slc39a5*^−/−^ mice to generate *Slc39a5*^-/-^; *Lepr*^-/-^. All studies were performed in both sexes. For HFFD study, ten-week-old mice were fed HFFD diet (46kcal% Fat, 30kcal% Fructose, TestDiet 5WK9) or control diet (TestDiet 58Y2) for 30 weeks. For NASH study, ten-week-old mice were fed NASH diet (40kcal% Fat, 20 kcal% Fructose and 2% Cholesterol, ResearchDiets D09100310) or control diet (ResearchDiets D09100304) for 40 weeks. All mice used in this study were housed in pathogen-free environment at Regeneron Pharmaceuticals Inc. animal research facility. Sterile water and chow were provided *ad libitum*. All experimental protocols and tissue harvesting procedures were performed with Regeneron Pharmaceuticals Inc., Institutional Animal Care and Use Committee (IACUC) approval. Data are presented as box plots with individual values. Statistical analysis was performed using one-way or two-way ANOVA, followed by post hoc Tukey’s tests. Statistical significance reported when p<0.05. Sample sizes, statistical test and significance are described in each figure legend.

Full Methods and any associated references are available in supplementary information.

## List of Supplementary Materials

Supplementary Methods Fig S1 to S16

Table S1 to S6

## Data Availability

All data produced in the present work are contained in the manuscript

## Acknowledgements

We wish to thank individuals that consented to be part of the Regeneron Genetics Center-Geisinger Health System DiscovEHR study. We would also like to acknowledge Kristy Neiman and William Poueymirou for their assistance with mouse colony management, and Suganthi Balasubramanian for insightful comments and input at the inception of the project, and Sergio Fazio for a critical reading of our manuscript.

## Author contributions

H.N., A.N.E. and S.M.C. designed the study. S.M.C., K.H., J.D., W.W. and H.N. generated and analyzed the data. H.N., C.V.H., M.A.R.F., B.Y., A.L., S.B., V.A., A.M., A.E.L., J.B., N.V., T.D., L.L., L.M., M.G.L., D.C., O.M., A.S., DiscovEHR and Regeneron Genetics Center performed the human genetics data collection and analysis. S.M.C., K.K., A.N.E. and H.N. wrote the manuscript.

---

**Suppl. Fig. 1.**
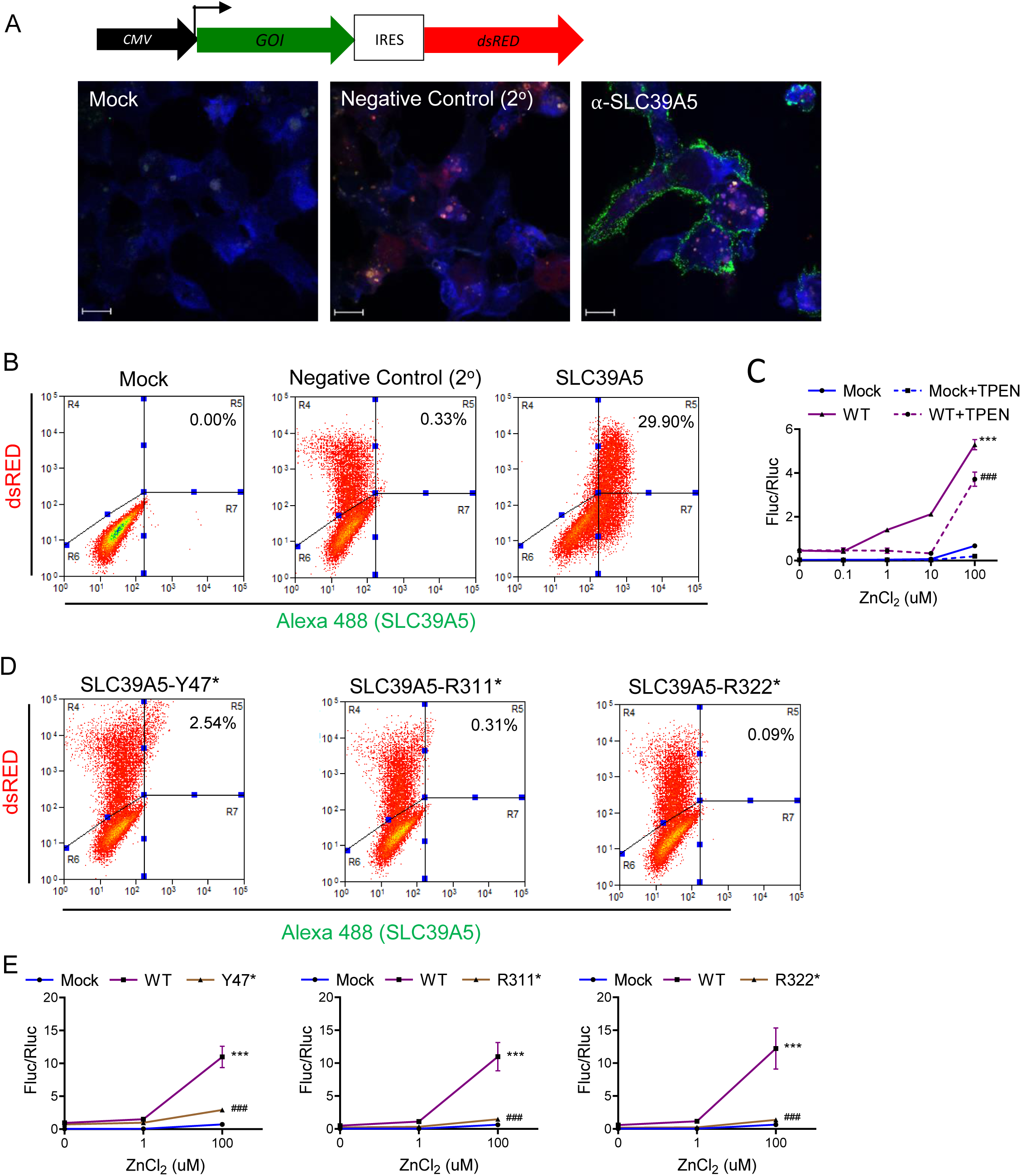
*SLC39A5* pLOF variants p.Y47*(c.141C>G), p.R311*(c.931C>T), and p.R322*(c.964C>T) encode for non-functional proteins. HEK293 cell transfected with expression constructs encoding SLC39A5 wild-type (WT), Y47*, R311* and R322* variants. (A, B) Immunostaining and FACS analysis demonstrating WT SLC39A5 localization to the cell surface. (C) Overexpression of WT SLC39A5 results in Zn^2+^ mediated *MRE* activation in a dose dependent manner, n=4. (D) FACS analyses demonstrating that cell surface expression of SLC39A5 Y47*, R311* and R322* muteins is markedly reduced. (E) Variants Y47*, R311* and R322* did not mediate Zn^2+^ induction of *MRE*, n=8, Statistical comparison to Mock and WT, respectively: ***P < 0.001, ^###^P < 0.001, two-way ANOVA with post hoc Tukey’s test. Metal regulatory element (*MRE*), firefly luciferase (Fluc), renilla luciferase (Rluc), cytomegalovirus (CMV), gene of interest (GOI), internal ribosome entry site (IRES).

**Suppl. Fig. 2.**
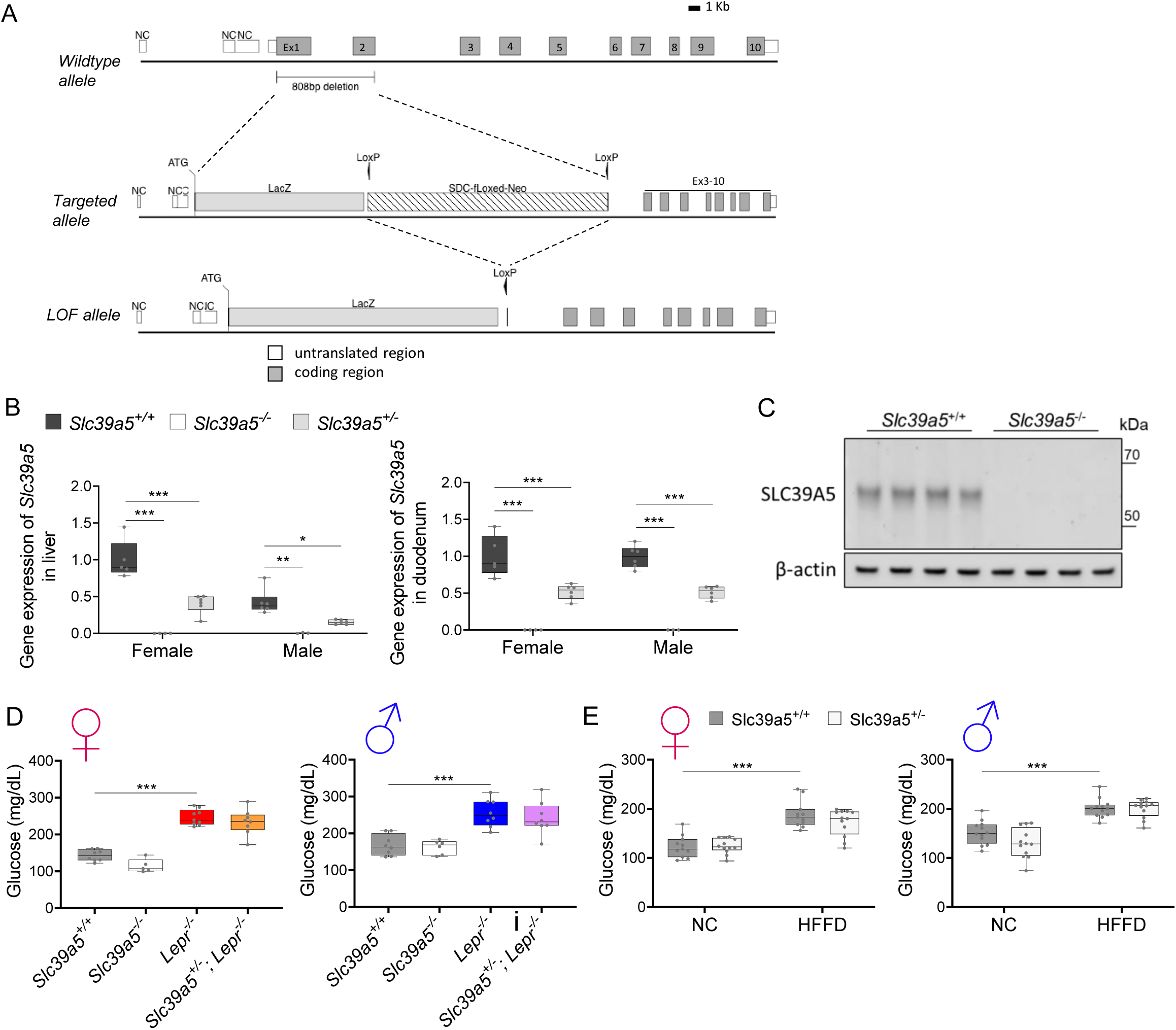
Generation and characterization of the *Slc39a5^-/-^* mice. (A) Schematic representation of the *Slc39a5* null allele. (B) *Slc39a5* gene expression in liver and duodenum of *Slc39a5*^-/-^ mice at 20 weeks of age, n=3-6. (C) Immunoblotting analyses demonstrating absence of SLC39A5 protein in liver of *Slc39a5*^-/-^ mice at 34 weeks of age. (D-E) Heterozygous loss of *Slc39a5* does not reduce fasting blood glucose in (D) *Lepr^-/-^* mice (at 20 weeks of age) and in (E) mice challenged with high fat high fructose diet (HFFD) for 18 weeks. *P < 0.05, **P < 0.01, ***P < 0.001, two-way ANOVA with post hoc Tukey’s test.

**Suppl. Fig. 3.**
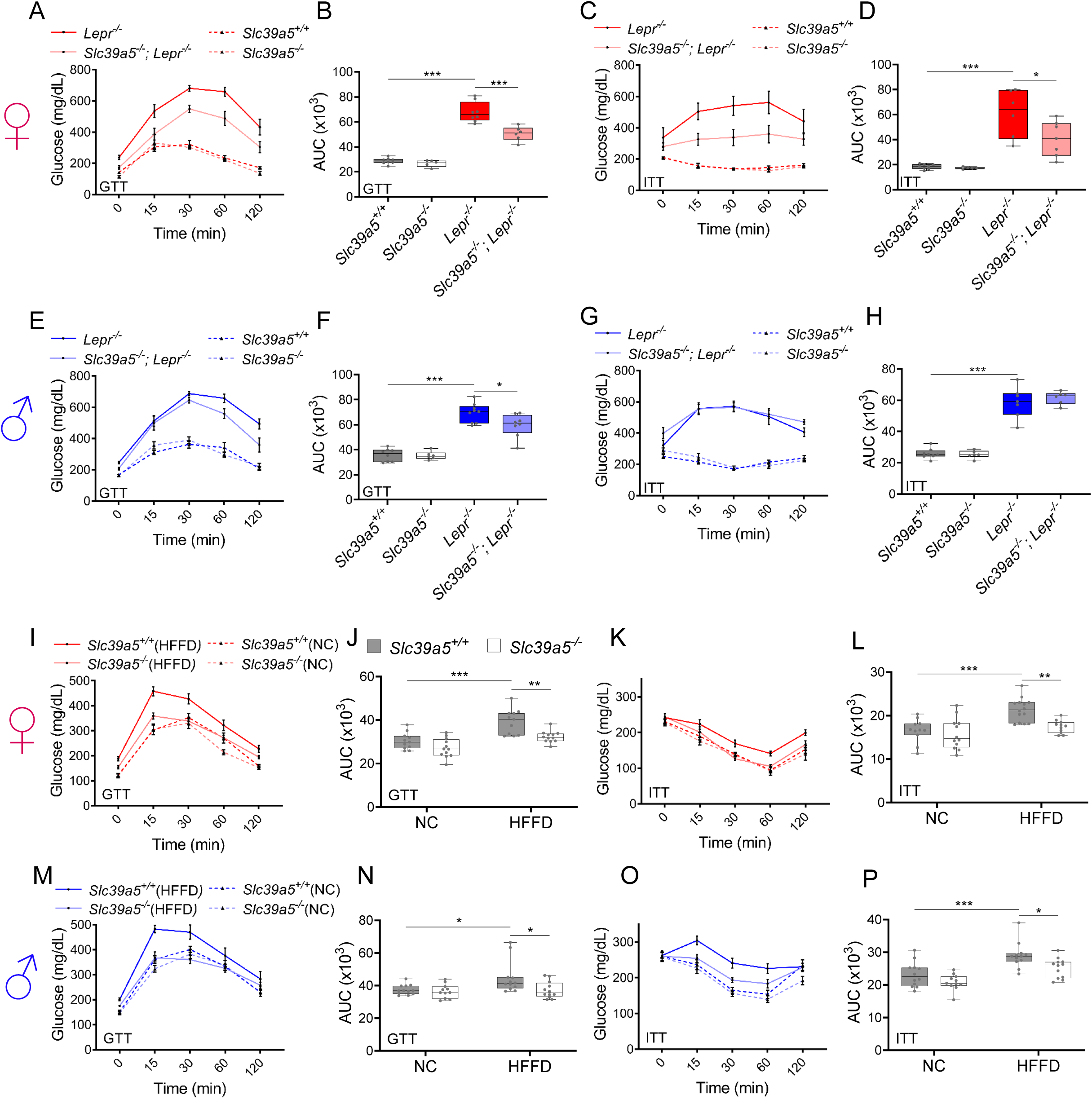
Loss of *Slc39a5* improves glycemic traits in *Lepr^-/-^* mice and in mice challenged with high fat high fructose diet (HFFD). Female **(A-D, I-L; ♀)** and Male **(E-H, M-P; ♂)** mice. (A-H) *Slc39a5^-/-^*; *Lepr*^-/-^ and corresponding control mice. (A-B, E-F) Oral glucose tolerance test (GTT) after 16 hour fasting, at 20 weeks. (C-D, G-H) Intraperitoneal insulin tolerance test (ITT), at 33 weeks. n=5-8. *P < 0.05, **P < 0.01, ***P < 0.001, one-way ANOVA with post hoc Tukey’s test. (I-P) HFFD mice. (I-J, M-N) GTT after 16 hours fasting, at 18 weeks. (K-L. O-P) ITT after 4 hour fasting, at 27 weeks. n=11-12. *P < 0.05, **P < 0.01, ***P < 0.001, two-way ANOVA with post hoc Tukey’s test. Area under curve (AUC). Numeric data is summarized in Suppl. Table 4 and 5.

**Suppl. Fig. 4.**
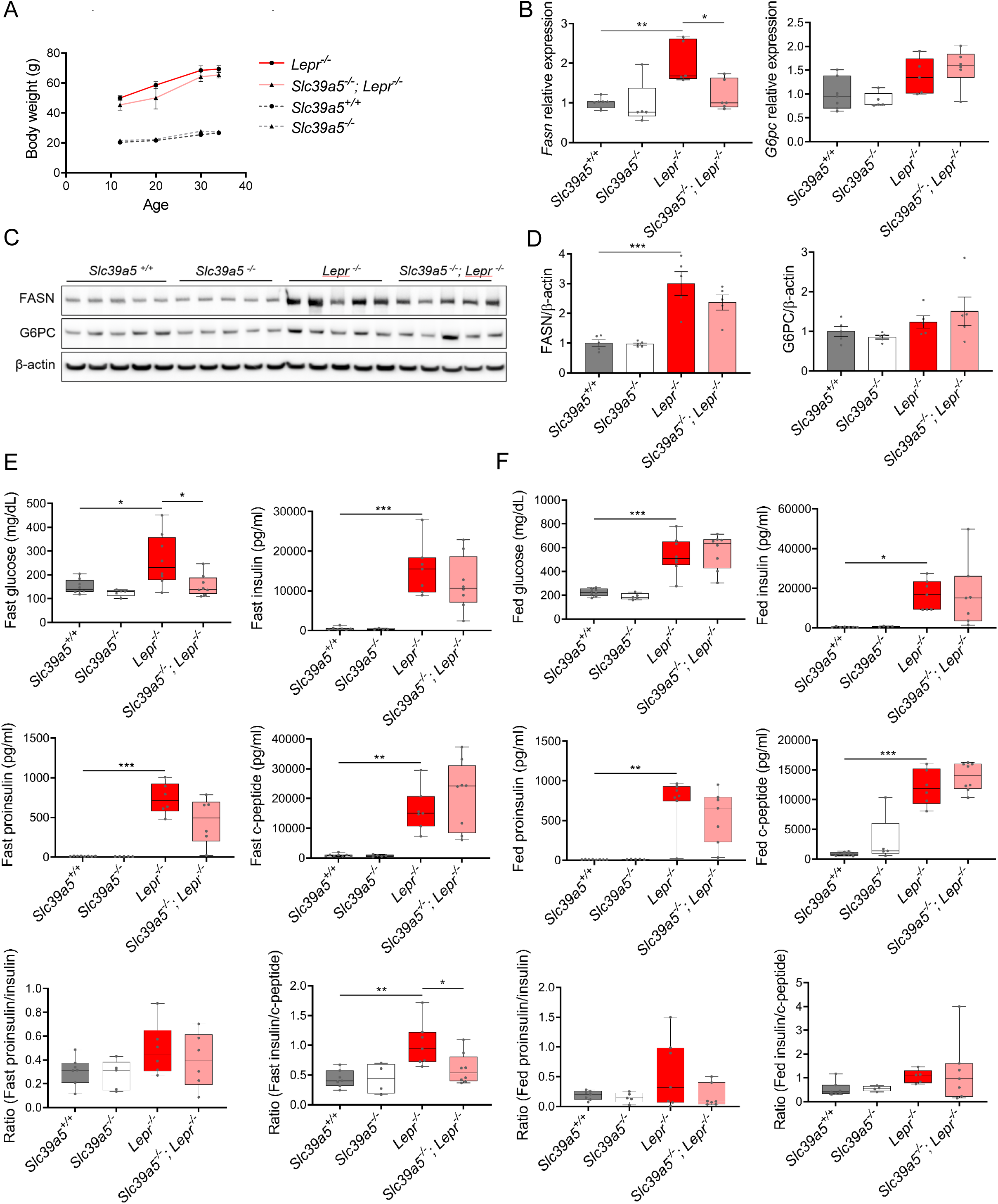
Metabolic profiling of female *Slc39a5^-/-^; Lepr^-/-^* mice. (A) Longitudinal body weight. (B-D) Analyses were done on explanted liver samples collected after 16 hour fasting at 34 weeks of age. (B) Hepatic expression of fatty acid synthase (*Fasn*) and glucose-6-phosphatase (*G6pc*). (C) Hepatic FASN and G6PC protein) Serum insulin profile upon fasting at 34 weeks of levels. (D) Densitometric analysis of hepatic FASN and G6PC. (F) Serum insulin profile in fed state at 32 weeks of age. *P < 0.05, **P < 0.01, ***P < 0.001, ANOVA with post hoc Tukey’s test.

**Suppl. Fig. 5.**
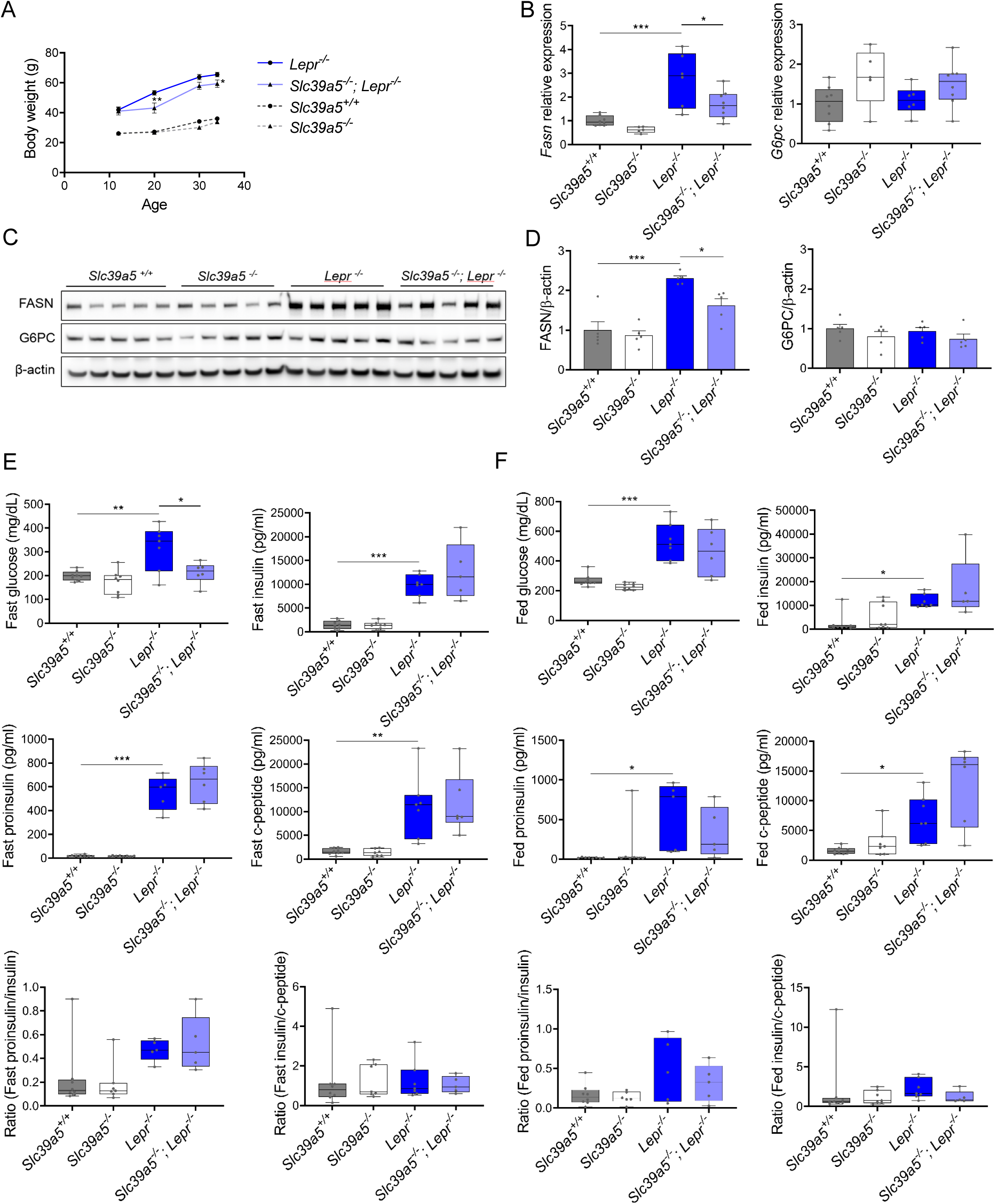
Loss of *Slc39a5* reduces hepatic fatty acid synthase expression but does not change insulin profile of male *Lepr^-/-^* mice. (A) Longitudinal body weight. (B-D) Analyses were done on explanted liver samples collected after 16 hour fasting at 34 weeks of age. (B) Hepatic expression of *Fasn* and *G6pc.* (C) Hepatic FASN and G6PC protein levels. (D) Densitometric analysis of hepatic FASN and G6PC. (E) Serum insulin profile upon fasting at 34 weeks of age. (F) Serum insulin profile in fed state at 32 weeks of age. *P < 0.05, **P < 0.01, ***P < 0.001, ANOVA with post hoc Tukey’s test.

**Suppl. Fig. 6.**
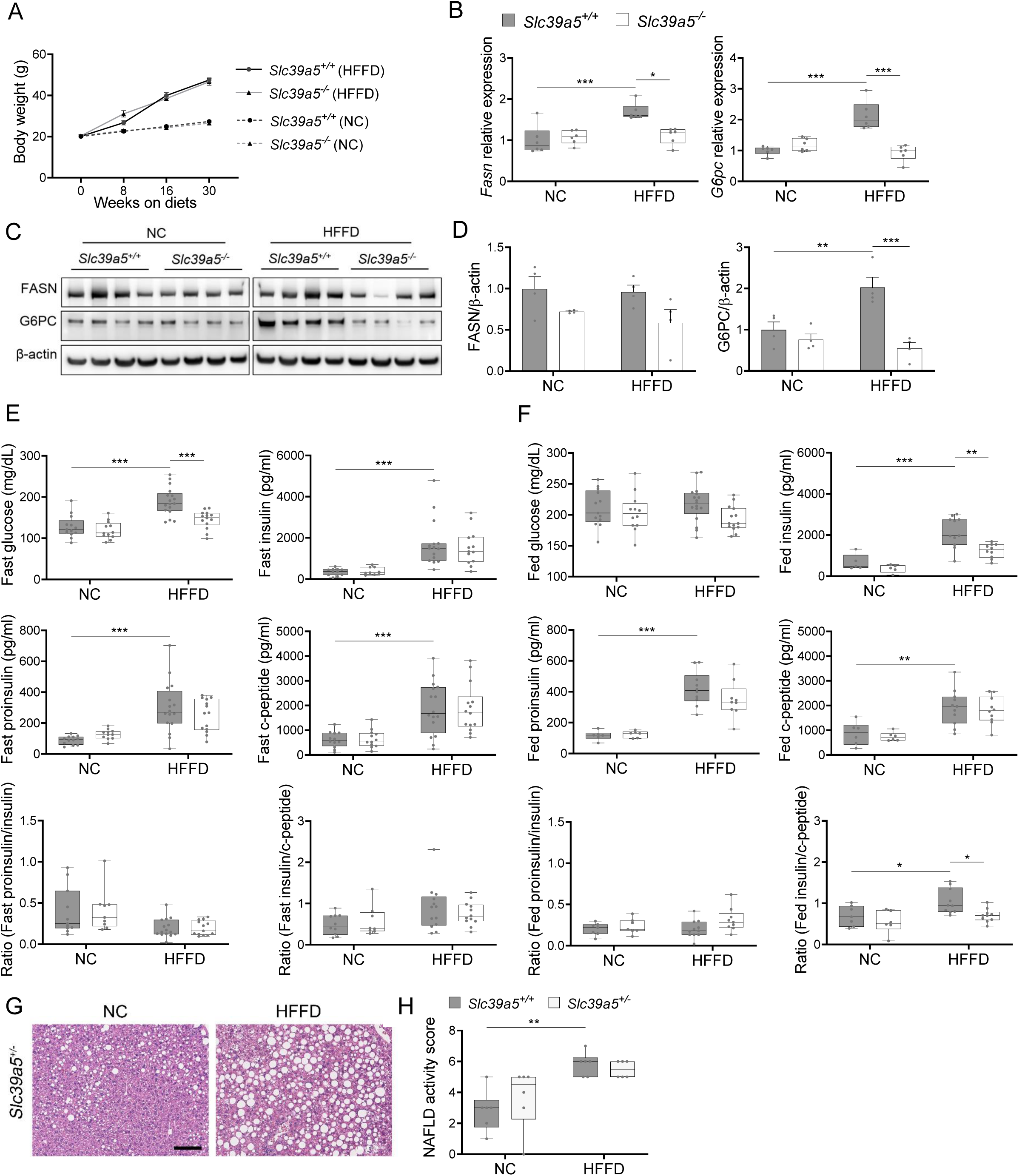
Loss of *Slc39a5* reduces hepatic fatty acid synthase expression but does not change insulin profile in female mice challenged with high fat high fructose diet (HFFD). (A) Longitudinal body weight during dietary intervention. (B-D) Analyses were done on explanted liver samples collected after 16 hour fasting in mice fed HFFD or NC for 30 weeks. (B) Hepatic expression of *Fasn* and *G6pc.* (C) Hepatic FASN and G6PC protein levels. (D) Densitometric analysis of hepatic FASN and G6PC. (E) Fasting serum insulin profile. (F) Fed serum insulin profile. (G) Representative images of *Slc39a5*^+/-^ livers stained with H&E. Scale bar, 100um. (H) NAFLD activity score. *P < 0.05, **P < 0.01, ***P < 0.001, two-way ANOVA with post hoc Tukey’s test.

**Suppl. Fig. 7.**
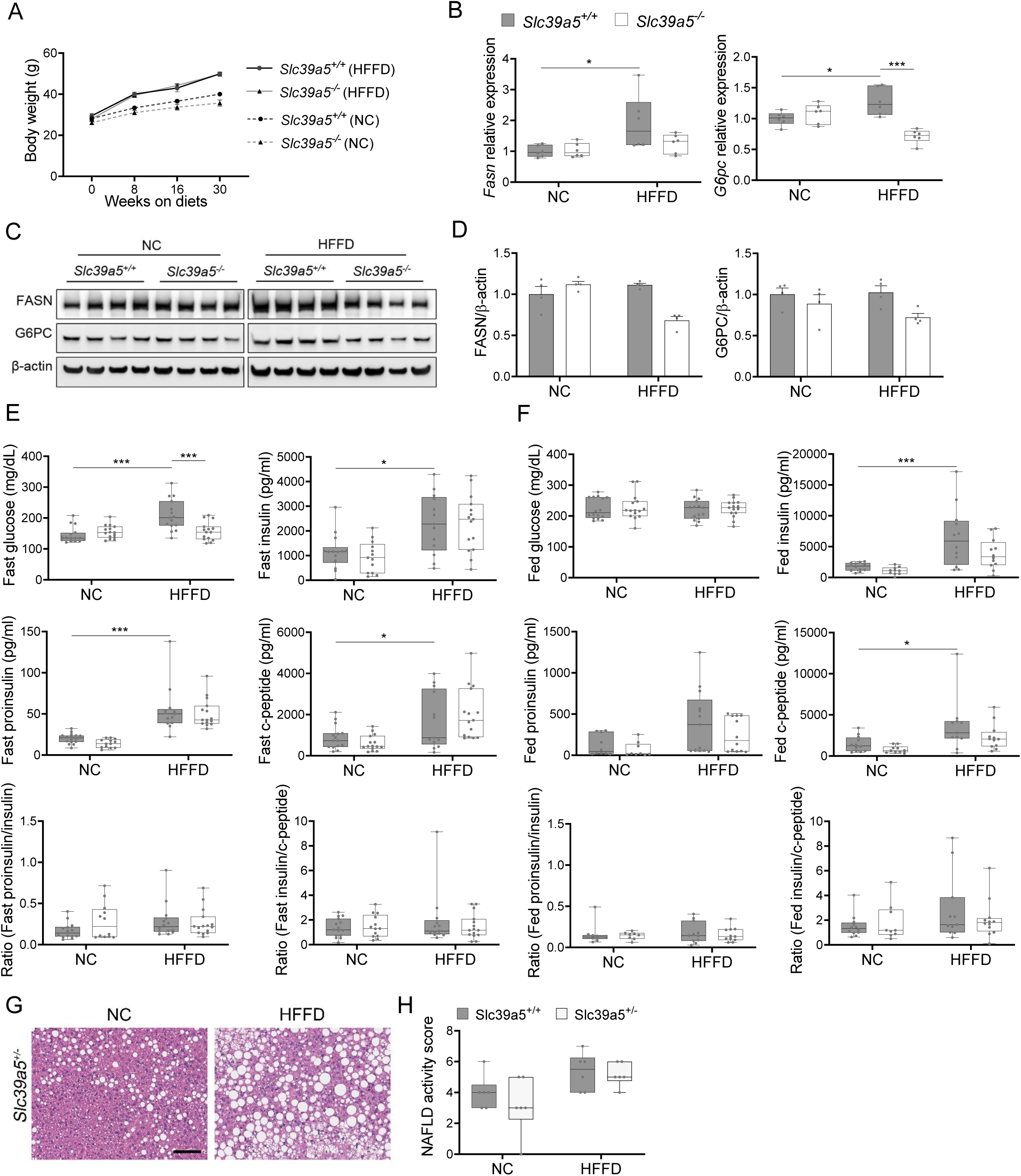
Loss of *Slc39a5* reduces hepatic fatty acid synthase expression but does not change insulin profile in male mice challenged with high fat high fructose diet (HFFD). (A) Longitudinal body weight during dietary intervention. (B-D) Analyses were done on explanted liver samples collected after 16 hour fasting in mice fed HFFD or NC for 30 weeks. (B) Hepatic expression of *Fasn* and *G6pc.* (C) Hepatic FASN and G6PC protein levels. (D) Densitometric analysis of hepatic FASN and G6PC. (E) Fasting serum insulin profile. (F) Fed serum insulin profile. (G) Representative images of *Slc39a5*^+/-^ livers stained with H&E. Scale bar, 100um. (H) NAFLD activity score. *P < 0.05, **P < 0.01, ***P < 0.001, two-way ANOVA with post hoc Tukey’s test.

**Suppl. Fig. 8.**
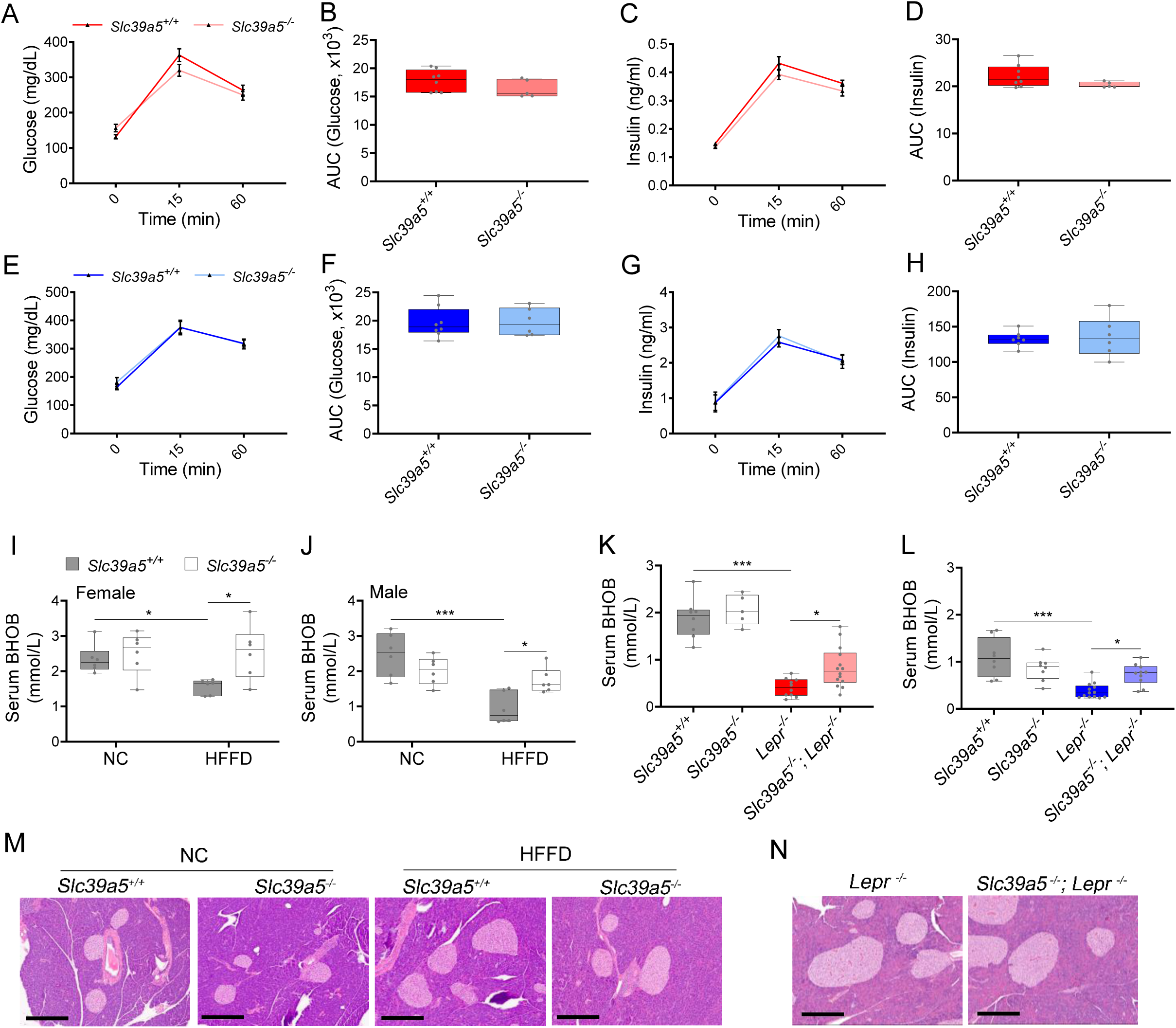
Additional data of glucose stimulated insulin secretion, serum BHOB and pancreas histology in mouse models. (A-H) Oral glucose tolerance test (GTT) was performed in *Slc39a5^-/-^* female (A-D) and male (E-H) mice after 16 hour fasting, at 15 weeks. Glucose (A-B, E-F) and insulin (C-D, G-H) levels were measured at 0, 15 and 60 mins. n=6-8. (I-J) Serum BHOB levels in female (I) and male (J) mice challenged with high fat high fructose diet (HFFD). *P < 0.05, ***P < 0.001, two-way ANOVA with post hoc Tukey’s test. (K-L) Serum BHOB levels in *Slc39a5^-/-^; Lepr^-/-^* female (K) and male (L) mice. *P < 0.05, ***P < 0.001, one-way ANOVA with post hoc Tukey’s test. (M-N) No overt morphological deficits in pancreas resulting from *Slc39a5* deficiency. Scale bar, 300µm

**Suppl. Fig. 9.**
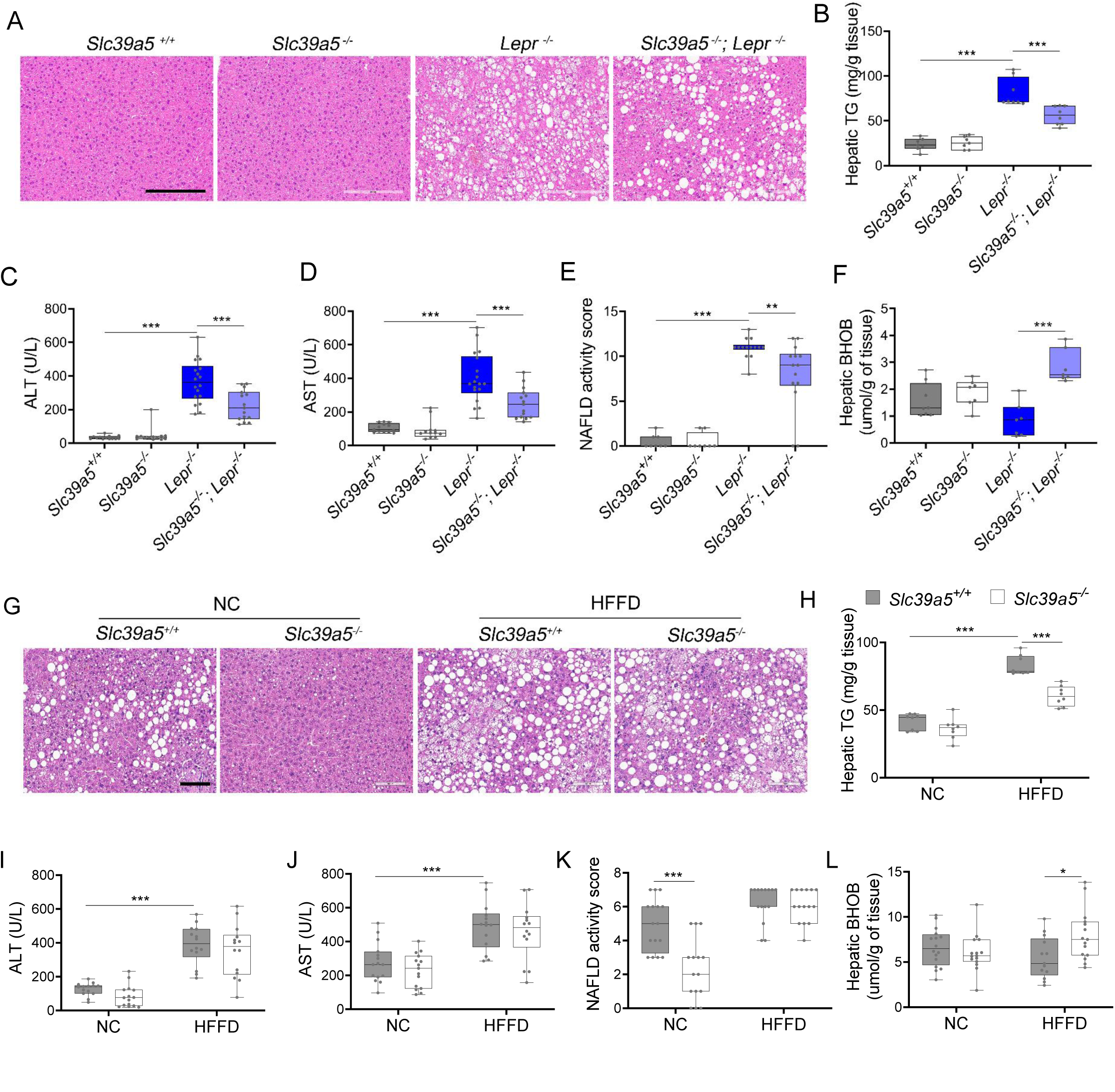
Loss of *Slc39a5* improves liver function and steatosis in *Lepr^-/-^* male mice and reduces hepatic triglyceride in male mice challenged with high fat high fructose diet (HFFD). *Slc39a5^-/-^; Lepr^-/-^* and corresponding control mice (A-F) were sacrificed after 16 hours fasting at 34 weeks of age. (G-L) *Slc39a5^-/-^* and corresponding control mice were fed HFFD or NC for 30 weeks and sacrificed after 16 hours fasting. (A, G) Representative images of livers stained with H&E. Scale bar, 200µm. (B, H) Hepatic triglyceride (TG) content in explanted liver samples at endpoint. (C, I) Serum ALT. (D, J) Serum AST. (E, K) NAFLD activity score, (F, L) Hepatic beta-hydroxybutyrate (BHOB). *P < 0.05, **P < 0.01, ***P < 0.001, *Slc39a5^-/-^; Lepr^-/-^* mice: one-way ANOVA with post hoc Tukey’s test, HFFD: two-way ANOVA with post hoc Tukey’s test. Numeric data is summarized in Suppl. Table 4 and 5.

**Suppl. Fig. 10.**
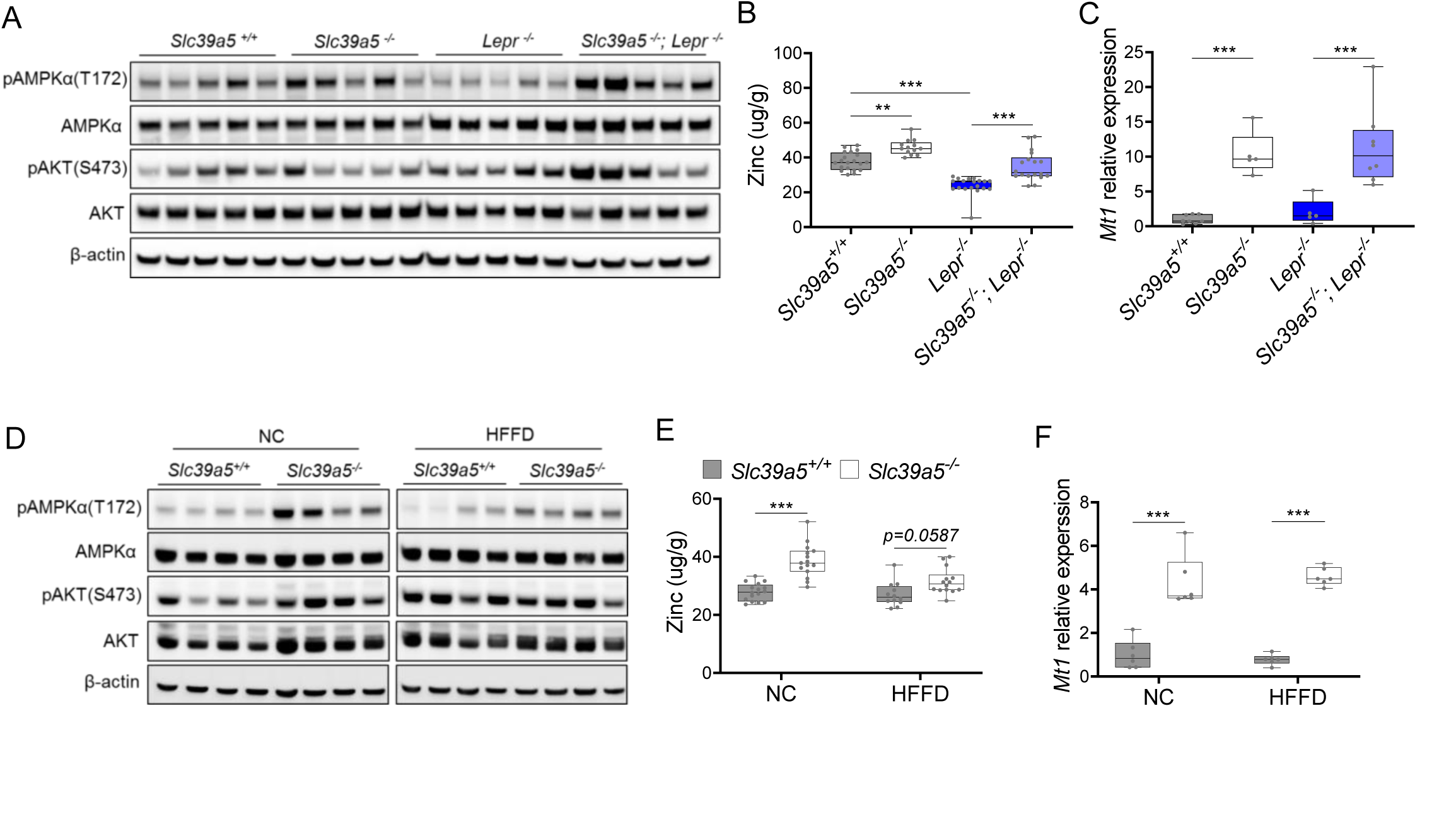
Loss of *Slc39a5* results in elevated hepatic zinc and activation of hepatic AMPK signaling in congenital and diet-induced obesity models. Analyses were done on explanted liver samples collected from male mice after 16 hour fasting at endpoint of congenital (A-C) and diet-induced obesity (D-F). (A, D) Immunoblot analysis of hepatic AMPK and AKT activation. (B, E) Hepatic zinc measurements (n=10-21). (C, F) Hepatic *Mt1* gene expression. *P < 0.05, **P < 0.01, ***P < 0.001, ANOVA with post hoc Tukey’s test.

**Suppl. Fig. 11.**
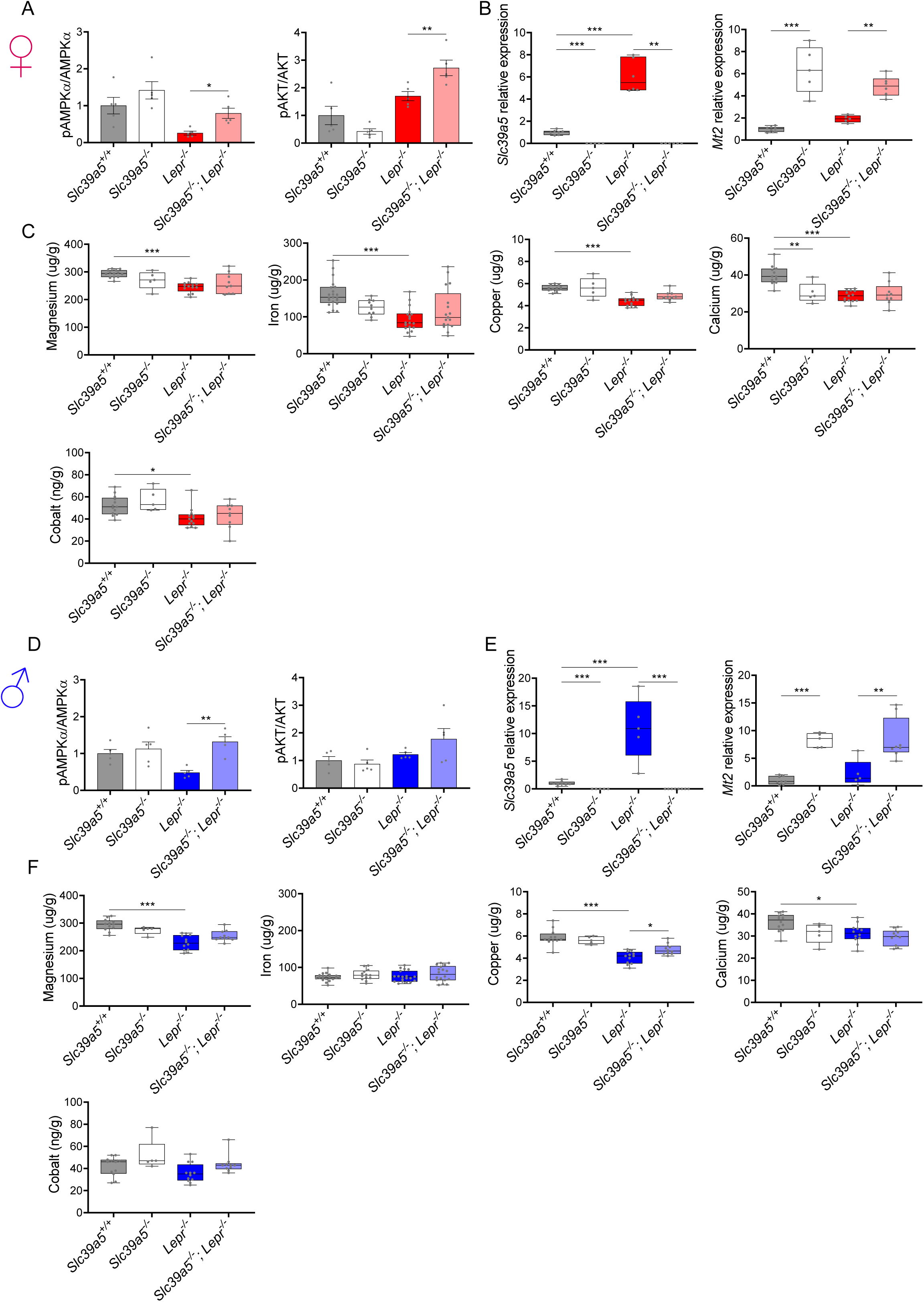
Loss of *Slc39a5* does not alter hepatic magnesium, iron, copper, calcium and cobalt levels in *Lepr^-/-^* mice. Female (A-C) and male (D-F) mice were examined at 34 weeks of age. (A, D) Densitometric analysis of hepatic AMPK and AKT signaling. (B, E) Hepatic expression of *Slc39a5* and *Mt2*. (C, F) Hepatic ion quantification by flame atomic absorption spectrometry. *P < 0.05, **P < 0.01, ***P < 0.001, ANOVA with post hoc Tukey’s test.

**Suppl. Fig. 12.**
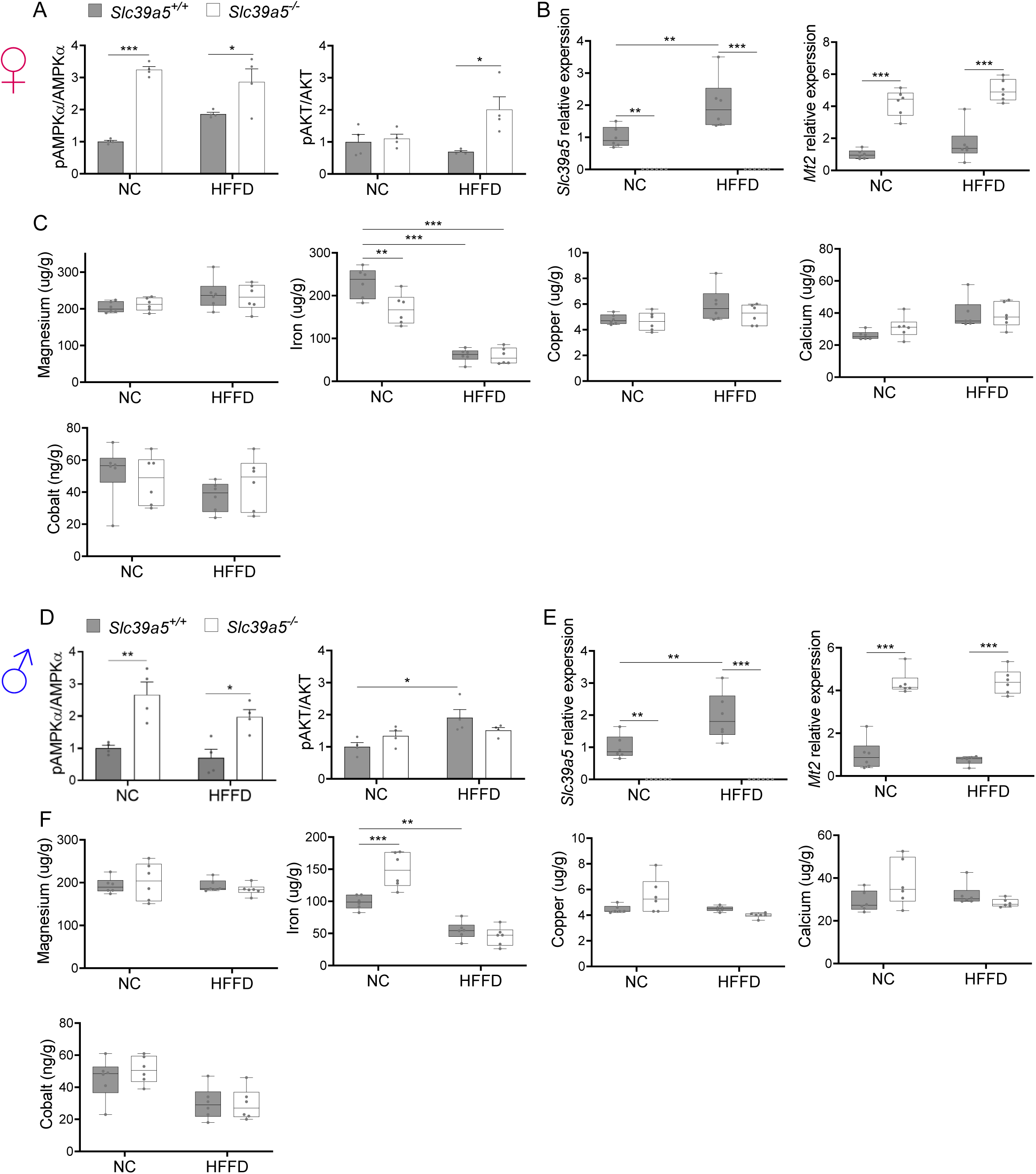
Loss of *Slc39a5* does not alter hepatic magnesium, iron, copper, calcium and cobalt levels in mice challenged with high fat high fructose diet (HFFD). Female (A-C) and male (D-F) mice were fed HFFD or NC for 30 weeks. (A, D) Densitometric analysis of hepatic AMPK and AKT. (B, E) Hepatic gene expression of *Slc39a5* and *Mt2*. (C, F) Hepatic ion quantification by flame atomic absorption spectrometry. *P < 0.05, **P < 0.01, ***P < 0.001, two-way ANOVA with post hoc Tukey’s test.

**Suppl. Fig. 13.**
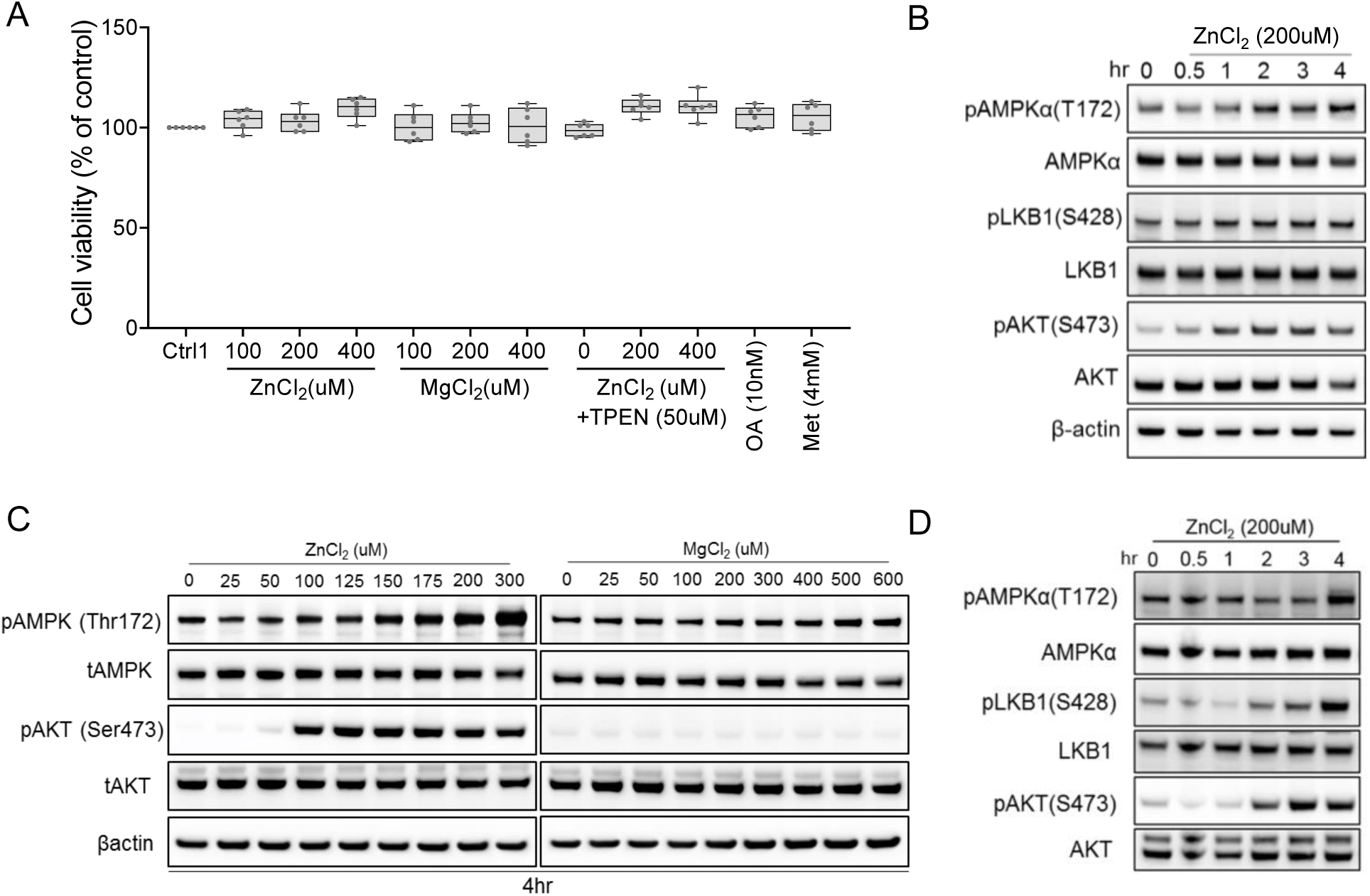
Zinc activates AMPK and AKT signaling in time-dependent and dose-dependent manner. (A) No differences in cell viability observed in human primary hepatocytes (after 4hr treatment) across different experimental groups. (B) Time-resolved (0-4hr) immunoblotting analyses of primary human hepatocytes treated with zinc chloride. (C) Immunoblots of HepG2 treated with zinc chloride (ZnCl_2_) and magnesium chloride (MgCl_2_). (D) Time-resolved (0-4hr) immunoblotting analyses of HepG2 treated with zinc chloride. Okadaic acid (OA), metformin (Met), N,N,N’,N’-Tetrakis(2-pyridylmethyl)ethylenediamine (TPEN).

**Suppl. Fig. 14.**
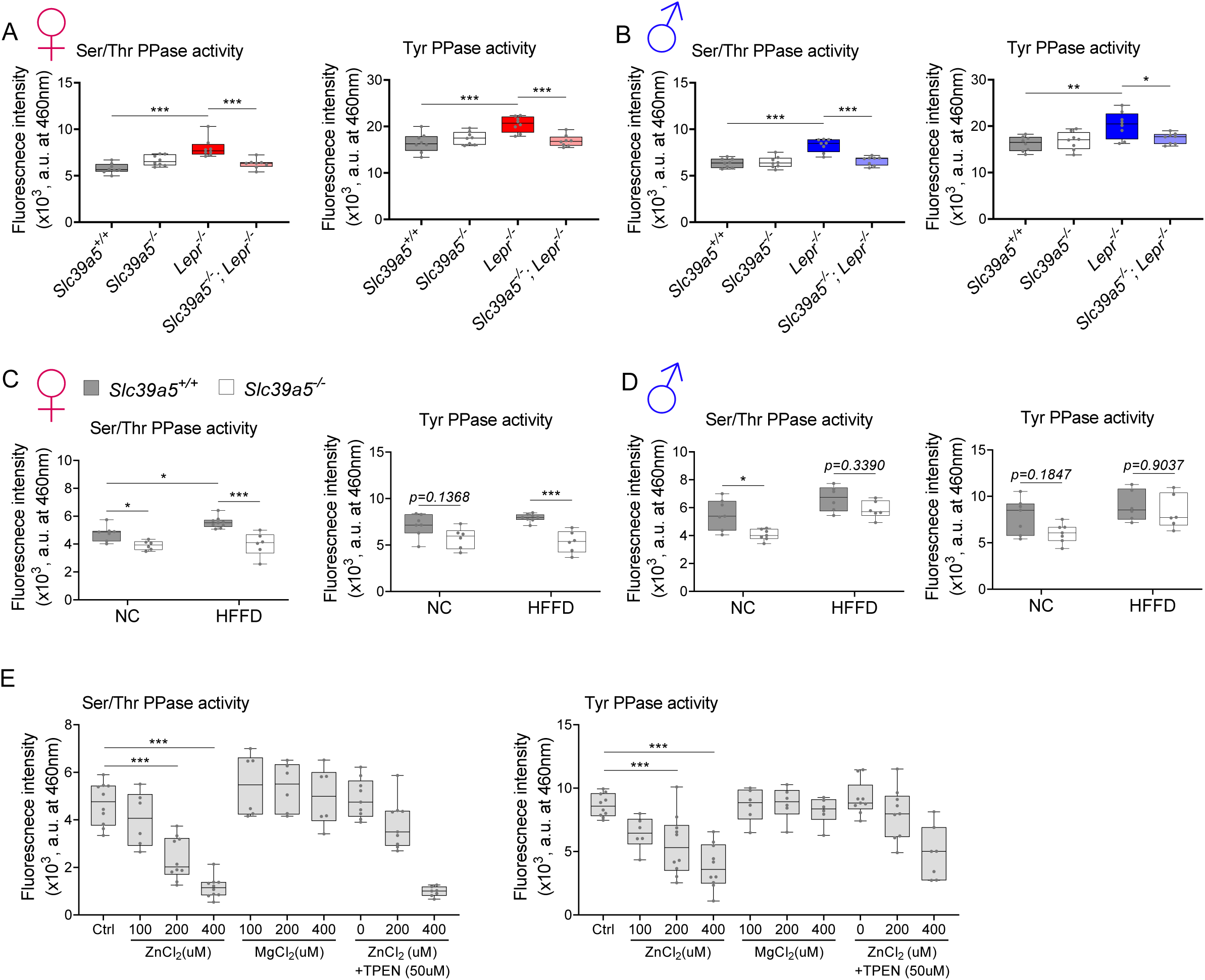
Elevated hepatic zinc results in reduced protein phosphatase activity. Analyses were done on explanted liver samples collected after 16 hour fast at endpoint of congenital obesity (A-B) and diet-induced obesity (C-D) challenges. Female (A, C) and Male (B, D) mice. (A-D) Ser/Thr and Tyr protein phosphatase activity. (E) Ser/Thr and Tyr protein phosphatase activity in primary human hepatocytes treated with zinc chloride (ZnCl_2_), magnesium chloride (MgCl_2_) and N,N,N’,N’-Tetrakis(2-pyridylmethyl)ethylenediamine (TPEN) for 4 hours. *P < 0.05, **P < 0.01, ***P < 0.001, ANOVA with post hoc Tukey’s test.

**Suppl. Fig. 15.**
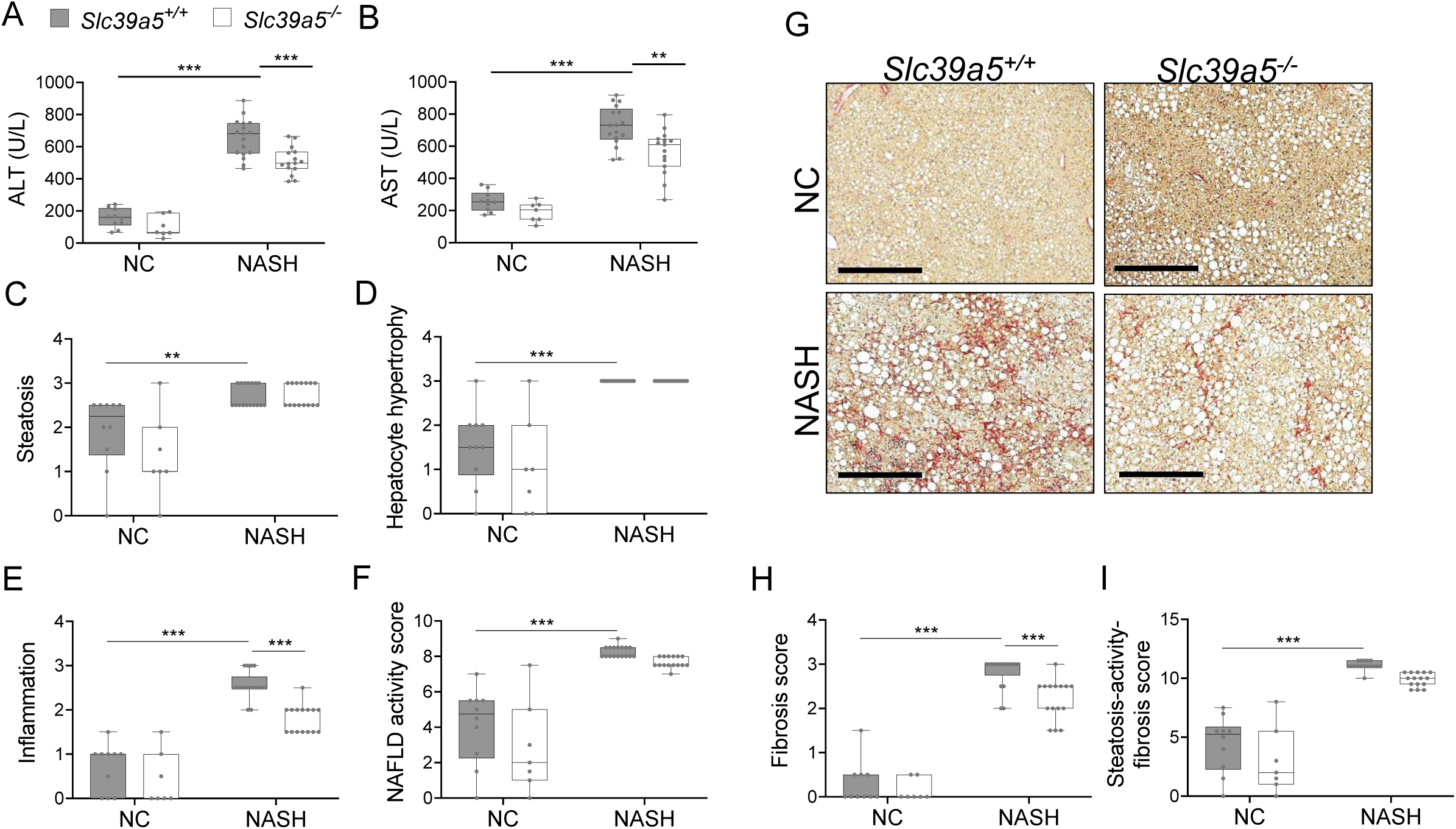
Loss of *Slc39a5* reduces hepatic inflammation and fibrosis in male mice challenged with diet-induced NASH. Mice were fed NASH diet or NC for 40 weeks and sacrificed after 16 hour fasting. (A-B) Loss of *Slc39a5* reduces serum ALT and AST levels (biomarkers of liver damage). (C-E) Histology scores for steatosis, hepatocyte hypertrophy, inflammation. (F) NAFLD activity score. (G-I) Loss of *Slc39a5* improves fibrosis in mice upon NASH dietary challenge. (G) Representative images of explanted livers sample stained with picrosirius red indicative of collagen deposition. Scale bar, 300µm. (H-I) Histology score for fibrosis and steatosis-activity-fibrosis score. n=7-10 (NC) and 15-17 (NASH), *P < 0.05, **P < 0.01, ***P < 0.001, two-way ANOVA with post hoc Tukey’s test. Numeric data is summarized in suppl. Table 6.

**Suppl. Fig. 16.**
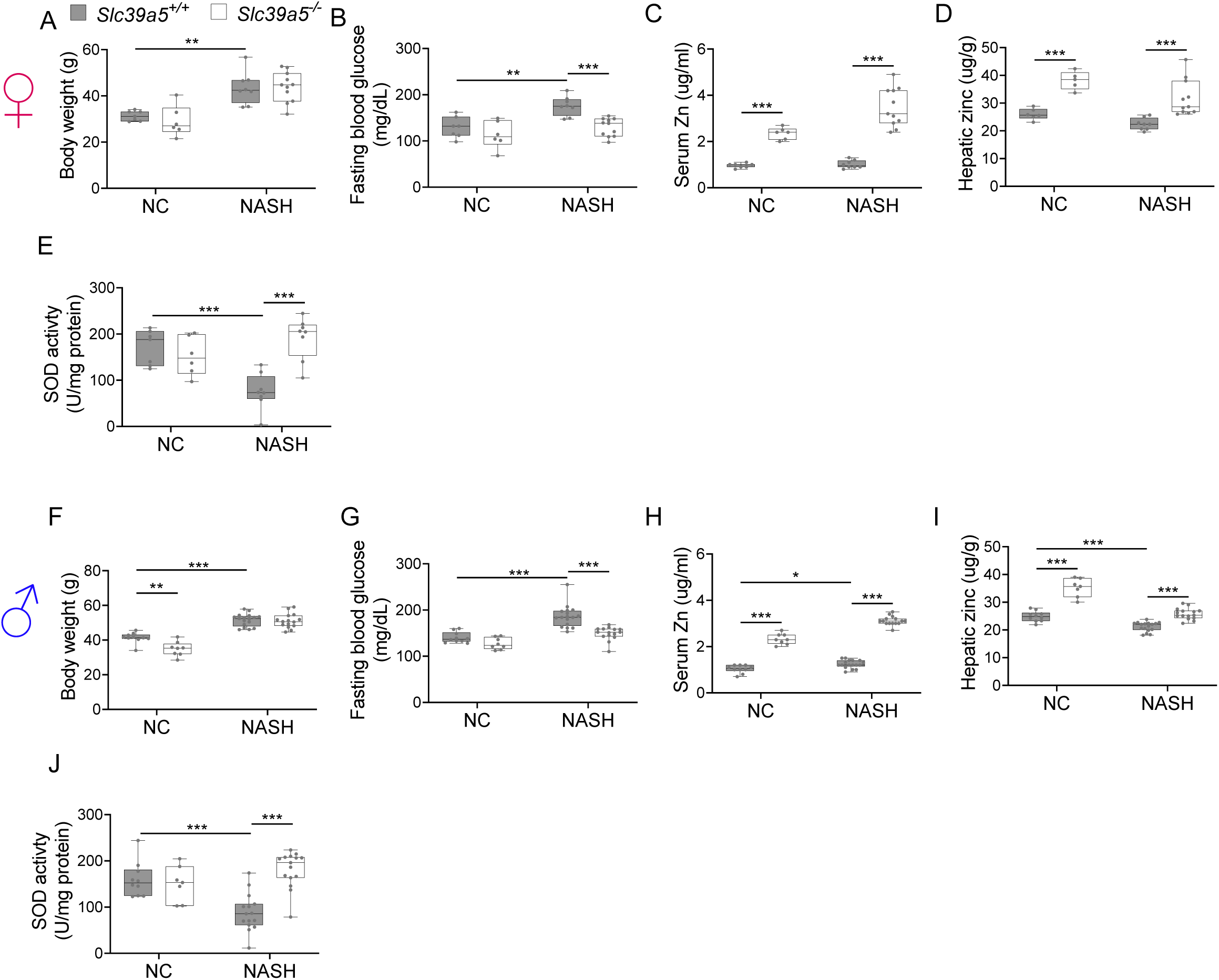
Loss of *Slc39a5* improves liver function in mice challenged with diet-induced NASH. Female (A-E) and Male (F-J) mice. (A, F) Body weight. (B, G) Fasting blood glucose. (C, H) Serum zinc. (D, I) Hepatic zinc. (E, J) Total hepatic SOD activity. n=6-10 (NC) and 8-17 (NASH), *P < 0.05, **P < 0.01, ***P < 0.001, two-way ANOVA with post hoc Tukey’s test.

**Suppl. Table 1.** Serum zinc and insulin profile assessment in the serum call back study. Serum zinc levels in *SLC39A5* heterozygous loss of function carriers are elevated by 12% as compared to age, sex, BMl-matched reference controls. Analyses of insulin production (insulin/c-peptide ratio), insulin clearance (proinsulin/insulin) and blood glucose in these samples demonstrated no differences based on genotype. Data represented in a graphical format in Fig. 1.

| Measure | Zinc |  | Insulin, pM |  | Proinsulin, pM |  | C-Peptide, pM |  | Proinsulin/Insulin |  | Insulin/C-Peptide |  | Glucose |  |
| --- | --- | --- | --- | --- | --- | --- | --- | --- | --- | --- | --- | --- | --- | --- |
| SLC39A5 | Ref | Het | Ref | Het | Ref | Het | Ref | Het | Ref | Het | Ref | Het | Ref | Het |
| Mean | 1.576 | 1.766 | 112.6 | 118.7 | 30.95 | 28.65 | 661.8 | 629.6 | 0.3222 | 0.2895 | 0.2331 | 0.2336 | 106.6 | 106.6 |
| Median | 1.513 | 1.744 | 70.5 | 82.05 | 18 | 16.7 | 475.1 | 480.6 | 0.258 | 0.239 | 0.145 | 0.154 | 103 | 106 |
| N | 248 | 90 | 246 | 86 | 253 | 91 | 253 | 91 | 246 | 86 | 247 | 86 | 247 | 87 |
| P values | 0.0024 |  | 0.682 |  | 0.5994 |  | 0.6449 |  | 0.2249 |  | 0.9924 |  | 0.9912 |  |

**Suppl. Table 2:**
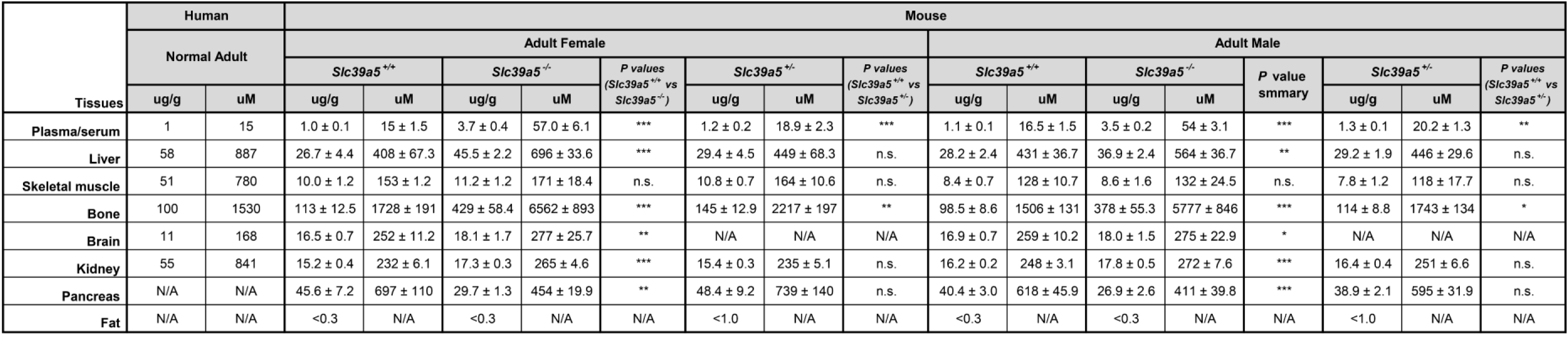
Tissue zinc content in human and mouse. Human data adapted from Jackson et. al^4^. *p<0.05; **p<0.01, ***p<0.001, not significant (n.s.), upaired t-test. Values represent mean ± SD.

**Suppl. Table 3:** No differences in serum chemistry profile of *Slc39a5^+I+^* and *Slc39a5^-/-^* mice. Serum chemistry analysis in adult mice (40 weeks of age, both sexes) demonstrated no differences in pancreatic amylase, renal function parameters (blood urea nitrogen, creatinine, total protein and uric acid) and electrolytes (chloride, potassium and sodium) or liver enzymes (alanine aminotransferase; ALT and aspartate aminotransferase; AST).

| Serum Chemistry<br>(40 weeks of age) | Female |  |  |  |  |  | Male |  |  |  |  |  |
| --- | --- | --- | --- | --- | --- | --- | --- | --- | --- | --- | --- | --- |
|  | Slc39a5<sup>+/+</sup> (n=6) |  | Slc39a5<sup>-/-</sup> (n=6) |  | Fold Change<br>(KO/WT) | P values | Slc39a5<sup>+/+</sup> (n=6) |  | Slc39a5<sup>-/-</sup> (n=6) |  | Fold Change<br>(KO/WT) | P values |
|  | Mean | SEM | Mean | SEM |  |  | Mean | SEM | Mean | SEM |  |  |
| ALT (U/l) | 43.00 | 5.99 | 35.90 | 7.17 | 0.83 | n.s. | 79.20 | 12.08 | 67.00 | 13.01 | 0.85 | n.s. |
| AST (U/l) | 147.00 | 21.49 | 180.60 | 53.50 | 1.23 | n.s. | 133.73 | 14.10 | 130.29 | 16.20 | 0.97 | n.s. |
| Albumin (g/dl) | 3.48 | 0.12 | 3.68 | 0.11 | 1.06 | n.s. | 4.13 | 0.09 | 3.98 | 0.17 | 0.96 | n.s. |
| Alkaline Phosphatase (U/l) | 113.50 | 10.09 | 132.17 | 9.12 | 1.16 | n.s. | 95.00 | 6.56 | 95.67 | 10.31 | 1.01 | n.s. |
| Alanine Transaminase (U/l) | 46.17 | 13.62 | 61.50 | 16.07 | 1.33 | n.s. | 129.00 | 16.43 | 53.83 | 17.84 | 0.42 | * |
| Aspartate Transaminase (U/l) | 169.50 | 24.64 | 192.17 | 37.17 | 1.13 | n.s. | 263.33 | 29.68 | 185.17 | 44.63 | 0.70 | n.s. |
| Amylase (U/l) | 3960.67 | 871.83 | 2864.20 | 368.35 | 0.72 | n.s. | 3317.40 | 272.77 | 2843.00 | 94.59 | 0.86 | n.s. |
| Blood Urea Nitrogen (mg/dl) | 20.50 | 0.89 | 21.83 | 0.65 | 1.07 | n.s. | 23.00 | 0.52 | 25.33 | 0.33 | 1.10 | * |
| Creatinine (mg/dl) | 0.13 | 0.02 | 0.16 | 0.02 | 1.22 | n.s. | 0.18 | 0.02 | 0.18 | 0.01 | 1.03 | n.s. |
| Total Protein (g/dl) | 5.23 | 0.20 | 5.77 | 0.10 | 1.10 | * | 6.23 | 0.12 | 5.83 | 0.22 | 0.94 | n.s. |
| Uric Acid (mg/dl) | 1.28 | 0.20 | 1.62 | 0.09 | 1.26 | n.s. | 2.75 | 0.30 | 2.25 | 0.27 | 0.82 | n.s. |
| Chloride (mmol/l) | 103.67 | 1.58 | 104.67 | 1.33 | 1.01 | n.s. | 106.50 | 0.67 | 106.50 | 2.05 | 1.00 | n.s. |
| Potassium (mmol/l) | 8.68 | 0.29 | 9.38 | 0.57 | 1.08 | n.s. | 10.00 | 0.46 | 11.35 | 0.43 | 1.14 | n.s. |
| Sodium (mmol/l) | 145.00 | 2.45 | 147.17 | 2.29 | 1.01 | n.s. | 149.67 | 1.33 | 147.00 | 2.49 | 0.98 | n.s. |
| Calcium (mg/dl) | 9.27 | 0.25 | 9.73 | 0.22 | 1.05 | n.s. | 10.07 | 0.16 | 9.93 | 0.22 | 0.99 | n.s. |
| Magnesium (mg/dl) | 2.57 | 0.13 | 2.93 | 0.11 | 1.14 | n.s. | 3.00 | 0.18 | 3.20 | 0.12 | 1.07 | n.s. |
| Iron (ug/dl) | 90.67 | 8.40 | 94.00 | 7.41 | 1.04 | n.s. | 114.00 | 6.88 | 99.33 | 4.74 | 0.87 | n.s. |

**Suppl. Table 4.** Summary statistics for the congenital obesity model. Loss of Slc39a5 improves glycemic traits and liver function in leptin-receptor *(Lepr)* deficient mice. Loss of Slc39a5 does not change insulin production (proinsulin/insulin), insulin clearance (insulin/c-peptide ratio). Data represented in a graphical format in Fig. 3, 4 and Suppl. Fig 5, 6.

|  | Sex | *Slc39a5*<sup>+/+</sup> | *Slc39a5*<sup>-/-</sup> | % change in *Slc39a5*<sup>-/-</sup> to *Slc39a5*<sup>+/+</sup> | P value | *Lepr*<sup>-/-</sup> | % change in *Lepr*<sup>-/-</sup> to *Slc39a5*<sup>+/+</sup> | P value | *Slc39a5*<sup>-/-</sup>; *Lepr*<sup>-/-</sup> | % change in *Slc39a5*<sup>-/-</sup>; *Lepr*<sup>-/-</sup> to *Lepr*<sup>-/-</sup> | P value |
| --- | --- | --- | --- | --- | --- | --- | --- | --- | --- | --- | --- |
| BW (g) at 34 weeks | F | 26.65 ± 0.92 | 27.30 ± 0.78 | 2% | n.s. | 69.32 ± 2.24 | 160% | *** | 65.36 ± 1.92 | -6% | n.s. |
|  | M | 36.03 ± 1.41 | 33.82 ± 1.24 | -6% | n.s. | 65.48 ± 1.49 | 82% | *** | 59.38 ± 2.48 | -9% | n.s. |
| Hepatic Zinc (ug/g) | F | 37.93 ± 1.17 | 55.56 ± 2.34 | 46% | *** | 25.53 ± 0.69 | -33% | *** | 32.04 ± 1.35 | 26% | ** |
|  | M | 37.79 ± 1.14 | 45.89 ± 1.22 | 21% | ** | 23.91 ± 1.19 | -37% | *** | 34.96 ± 1.98 | 46% | *** |
| Fasting Blood Glucose (mg/dL) | F | 142.88 ± 5.46 | 114.20 ± 8.19 | -20% | n.s. | 237.63 ± 13.54 | 66% | *** | 173.00 ± 11.65 | -27% | ** |
|  | M | 167.75 ± 10.00 | 162.50 ± 7.89 | -3% | n.s. | 245.78 ± 8.60 | 47% | *** | 207.75 ± 8.08 | -15% | * |
| GTT AUC (x10<sup>3</sup>) | F | 28.63 ± 0.86 | 26.70 ± 1.22 | -7% | n.s. | 67.74 ± 2.86 | 137% | *** | 50.55 ± 2.34 | -25% | *** |
|  | M | 35.75 ± 1.84 | 35.36 ± 1.34 | -1% | n.s. | 69.38 ± 2.54 | 94% | *** | 59.48 ± 3.27 | -14% | * |
| ITT AUC (x10<sup>3</sup>) | F | 18.38 ± 0.71 | 17.25 ± 0.37 | -6% | n.s. | 60.83 ± 7.71 | 231% | *** | 40.66 ± 5.27 | -33% | * |
|  | M | 25.79 ± 1.17 | 25.30 ± 1.04 | -2% | n.s. | 58.55 ± 3.74 | 127% | *** | 61.60 ± 1.66 | 5% | n.s. |
| HOMA-IR | F | 7.06 ± 1.94 | 2.16 ± 0.67 | -69% | n.s. | 229.79 ± 29.97 | 3153% | *** | 60.32 ± 16.58 | -74% | *** |
|  | M | 13.44 ± 3.10 | 15.26 ± 3.54 | 14% | n.s. | 228.10 ± 26.37 | 1597% | *** | 138.12 ± 14.78 | -39% | ** |
| Ratio (proinsulin/insulin) | F | 0.19 ± 0.03 | 0.18 ± 0.03 | -4% | n.s. | 0.55 ± 0.23 | 196% | n.s. | 0.27 ± 0.11 | -51% | n.s. |
|  | M | 0.16 ± 0.05 | 0.13 ± 0.03 | -20% | n.s. | 0.48 ± 0.18 | 190% | n.s. | 0.29 ± 0.13 | -40% | n.s. |
| Ratio (insulin/c-peptide) | F | 0.53 ± 0.12 | 0.55 ± 0.05 | 2% | n.s. | 1.06 ± 0.13 | 99% | n.s. | 1.27 ± 0.50 | 19% | n.s. |
|  | M | 2.08 ± 1.45 | 1.15 ± 0.33 | -45% | n.s. | 2.20 ± 0.48 | 6% | n.s. | 1.17 ± 0.35 | -47% | n.s. |
| ALT (U/L) | F | 31.64 ± 3.43 | 51.90 ± 11.25 | 64% | n.s. | 393.53 ± 31.59 | 1144% | *** | 200.94 ± 22.66 | -49% | *** |
|  | M | 34.46 ± 2.84 | 46.08 ± 13.00 | 34% | n.s. | 365.11 ± 29.73 | 959% | *** | 223.27 ± 22.89 | -39% | *** |
| AST (U/L) | F | 136.91 ± 19.22 | 150.00 ± 14.57 | 10% | n.s. | 514.47 ± 74.33 | 276% | *** | 264.06 ± 22.23 | -49% | *** |
|  | M | 104.92 ± 7.13 | 93.83 ± 16.83 | -11% | n.s. | 400.72 ± 35.12 | 282% | *** | 251.07 ± 23.50 | -37% | *** |
| NAFLD activity score | F | 1.20 ± 0.36 | 1.50 ± 0.22 | 25% | n.s. | 9.25 ± 0.18 | 671% | *** | 7.25 ± 0.58 | -22% | ** |
|  | M | 1.25 ± 0.25 | 1.25 ± 0.25 | 0% | n.s. | 9.36 ± 0.13 | 649% | *** | 7.15 ± 0.83 | -24% | * |
| Hepatic TG (mg/g) | F | 14.73 ± 1.53 | 15.82 ± 3.28 | 7% | n.s. | 67.11 ± 3.01 | 356% | *** | 29.24 ± 3.29 | -56% | *** |
|  | M | 23.64 ± 2.61 | 25.37 ± 2.63 | 7% | n.s. | 81.12 ± 5.56 | 243% | *** | 55.98 ± 3.65 | -31% | *** |

**Suppl. Table 5.** Summary statistics for the diet-induced obesity model. Loss of *Slc39a5* improves glycemic traits and liver function in mice upon a high fat high fructose diet (HFFD) dietary challenge. Moreover, loss of *Slc39a5* does not change insulin production (proinsulin/insulin), insulin clearance (insulin/c-peptide ratio). Data represented in a graphical format in Fig. 3, 4 and Suppl. Fig 7, 8.

|  | Sex | *Slc39a5*<sup>+/+</sup> (NC) | *Slc39a5*<sup>-/-</sup> (NC) | % change in *Slc39a5*<sup>-/-</sup> (NC) to *Slc39a5*<sup>+/+</sup> (NC) | P value | *Slc39a5*<sup>+/+</sup> (HFFD) | % change in *Slc39a5*<sup>+/+</sup> (HFFD) to *Slc39a5*<sup>+/+</sup> (NC) | P value | *Slc39a5*<sup>-/-</sup> (HFFD) | % change in *Slc39a5*<sup>-/-</sup> (HFFD) to *Slc39a5*<sup>+/+</sup> (HFFD) | P value |
| --- | --- | --- | --- | --- | --- | --- | --- | --- | --- | --- | --- |
| BW (g), diet for 30 weeks | F | 27.32 ± 0.61 | 26.53 ± 0.87 | -3% | n.s. | 47.51 ± 1.10 | 74% | *** | 46.65 ± 1.66 | -2% | n.s. |
|  | M | 40.03 ± 0.42 | 35.70 ± 1.48 | -11% | ** | 49.95 ± 0.91 | 25% | *** | 49.74 ± 0.79 | 0% | n.s. |
| Hepatic Zinc (ug/g) | F | 30.39 ± 1.38 | 43.82 ± 1.92 | 44% | ** | 32.73 ± 1.22 | 8% | n.s. | 47.08 ± 3.44 | 44% | *** |
|  | M | 27.91 ± 0.76 | 38.60 ± 1.49 | 38% | *** | 27.07 ± 1.20 | -3% | n.s. | 31.59 ± 1.19 | 17% | n.s. |
| Fasting Blood Glucose (mg/dL) | F | 122.82 ± 6.73 | 119.50 ± 8.04 | -3% | n.s. | 188.36 ± 8.24 | 53% | *** | 154.67 ± 7.28 | -18% | * |
|  | M | 151.50 ± 6.85 | 143.91 ± 6.35 | -5% | n.s. | 201.08 ± 5.59 | 33% | *** | 175.83 ± 3.52 | -13% | * |
| GTT AUC (x10<sup>3</sup>) | F | 30.33 ± 1.16 | 27.14 ± 1.26 | -11% | n.s. | 39.20 ± 1.74 | 29% | *** | 32.26 ± 0.76 | -18% | ** |
|  | M | 37.44 ± 0.88 | 36.59 ± 1.28 | -2% | n.s. | 44.70 ± 2.85 | 19% | * | 37.33 ± 1.48 | -16% | * |
| ITT AUC (x10<sup>3</sup>) | F | 16.56 ± 0.81 | 15.80 ± 1.03 | -5% | n.s. | 21.32 ± 0.72 | 29% | *** | 17.43 ± 0.45 | -18% | ** |
|  | M | 23.18 ± 1.10 | 20.87 ± 0.73 | -10% | n.s. | 29.05 ± 1.14 | 25% | *** | 25.22 ± 0.87 | -13% | * |
| HOMA-IR | F | 2.42 ± 0.38 | 1.88 ± 0.36 | -22% | n.s. | 16.20 ± 2.61 | 569% | *** | 10.27 ± 1.24 | -37% | * |
|  | M | 12.14 ± 2.64 | 7.65 ± 1.59 | -37% | n.s. | 31.69 ± 3.93 | 161% | *** | 17.97 ± 2.07 | -43% | ** |
| Ratio (proinsulin/insulin) | F | 0.20 ± 0.03 | 0.23 ± 0.03 | 17% | n.s. | 0.20 ± 0.03 | -1% | n.s. | 0.31 ± 0.05 | 57% | n.s. |
|  | M | 0.16 ± 0.04 | 0.14 ± 0.02 | -10% | n.s. | 0.18 ± 0.04 | 13% | n.s. | 0.16 ± 0.03 | -13% | n.s. |
| Ratio (insulin/c-peptide) | F | 0.70 ± 0.09 | 0.54 ± 0.11 | -22% | n.s. | 1.04 ± 0.10 | 50% | * | 0.70 ± 0.05 | -32% | * |
|  | M | 1.52 ± 0.26 | 1.82 ± 0.50 | 20% | n.s. | 2.89 ± 0.82 | 90% | n.s. | 2.06 ± 0.45 | -29% | n.s. |
| ALT (U/L) | F | 49.46 ± 7.98 | 60.83 ± 10.80 | 23% | n.s. | 241.94 ± 13.84 | 389% | *** | 148.71 ± 13.89 | -39% | *** |
|  | M | 126.71 ± 10.10 | 84.00 ± 16.84 | -34% | n.s. | 388.14 ± 30.94 | 206% | *** | 353.64 ± 41.42 | -9% | n.s. |
| AST (U/L) | F | 173.50 ± 15.07 | 216.83 ± 33.49 | 25% | n.s. | 449.71 ± 30.60 | 159% | *** | 370.79 ± 34.35 | -18% | n.s. |
|  | M | 276.40 ± 29.93 | 228.47 ± 25.98 | -17% | n.s. | 489.79 ± 37.65 | 77% | *** | 458.36 ± 44.22 | -6% | n.s. |
| NAFLD activity score | F | 4.50 ± 0.72 | 2.50 ± 0.72 | -44% | n.s. | 8.33 ± 0.33 | 85% | ** | 3.33 ± 0.71 | -60% | *** |
|  | M | 5.50 ± 0.43 | 1.83 ± 0.75 | -67% | ** | 7.83 ± 0.70 | 42% | n.s. | 8.67 ± 0.80 | 11% | n.s. |
| Hepatic TG (mg/g) | F | 23.42 ± 2.64 | 22.36 ± 1.80 | -5% | n.s. | 43.53 ± 1.09 | 86% | *** | 28.42 ± 1.74 | -35% | *** |
|  | M | 41.52 ± 2.11 | 36.29 ± 2.76 | -13% | n.s. | 83.01 ± 2.59 | 100% | *** | 60.41 ± 2.60 | -27% | *** |

**Suppl. Table 6.**
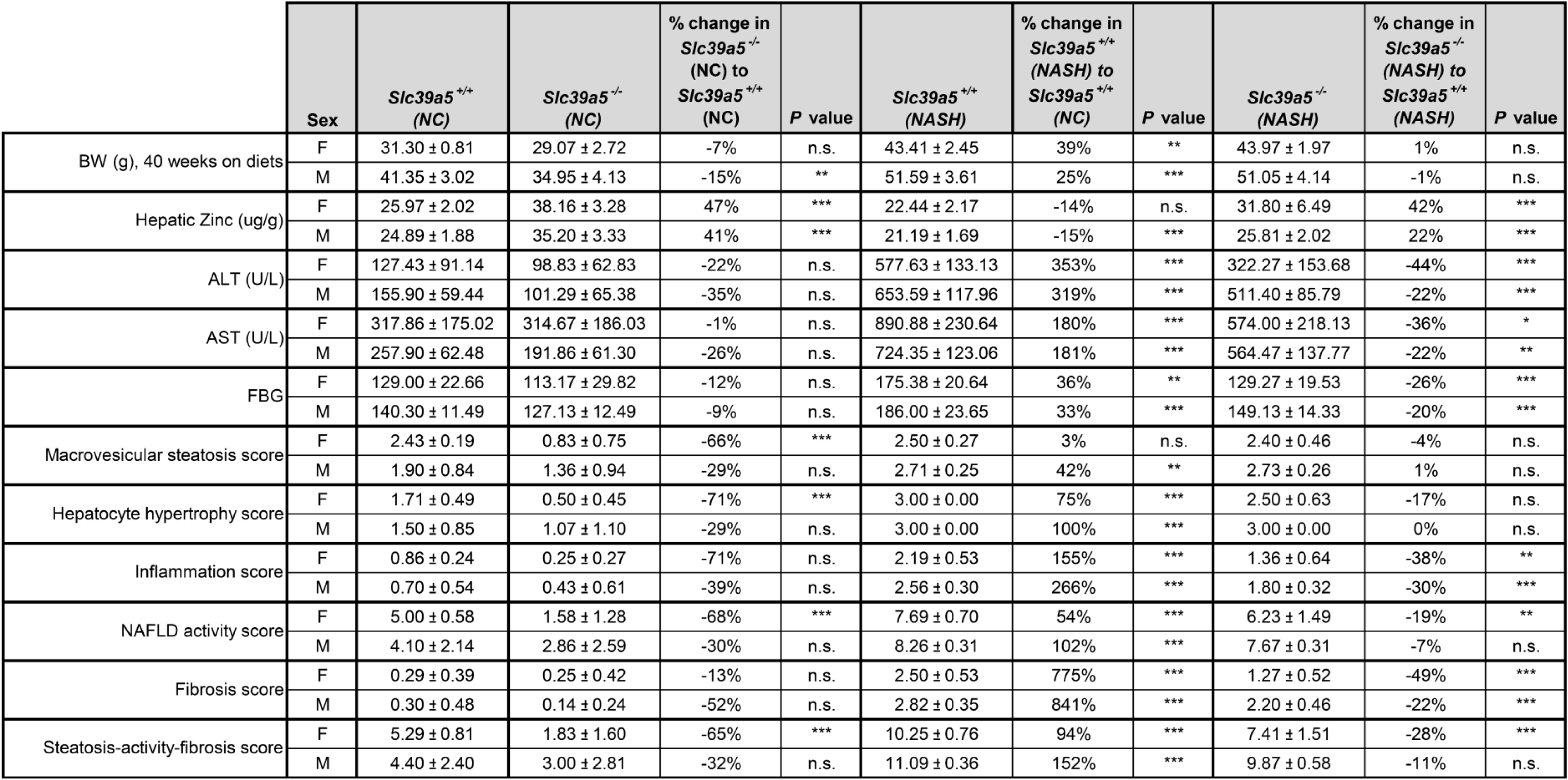
Summary statistics for the diet-induced NASH model. Loss of Slc39a5 improves hepatic inflammation and fibrosis in both female and male mice challenged with diet-induced NASH. Data represented in a graphical format in Fig. 6 and Suppl. Fig 14.

## Supplementary Methods

### Human Genetic Studies and Phenotyping

The Geisinger Health System DiscovEHR study is a hospital-based cohort of patients of the GHS, a large healthcare provision network in Central and Eastern Pennsylvania, United States. More than 200,000 health system participants have been enrolled and >145,000 have had exome sequencing performed by Regeneron Genetics Center. Type 2 diabetes (T2D) cases in DiscovEHR were defined as individuals with an ICD9 (code 250) or ICD10 (code E11) code for T2D, and either a median HbA1c value greater than or equal to 6.5%, or with a prescription for any diabetic medication. Individuals were excluded from the case pool if they had both an ICD10 code for type 1 diabetes (T1D; code E10) and if they did not have a prescription for any oral hypoglycemic medication. Controls were defined as individuals with no ICD codes for T1D or T2D, a median HbA1c value of less than 5.7%, and with no record of a prescription for any diabetic medication.

The UK Biobank is a prospective biomedical study of ∼500,00 adults from across the UK, including extensive phenotype measures and genomic data. T2D in UK Biobank was defined in line with a previously reported definition in this cohort^1, 2^. The UKB self-reported data were used to identify individuals with “probable type 2 diabetes”, “possible type 2 diabetes”, “probable type 1 diabetes” or “possible type 1 diabetes”, using a previously published algorithm^1^. T2D cases were defined as individuals with “probable type 2 diabetes” on self-report, or an ICD10 code E11 for T2D. Individuals were excluded from the analysis if they had “probable type 1 diabetes”, “possible type 1 diabetes”, or ICD10 code E10, for T1D.

The BioMe study (SINAI) is a highly diverse electronic health record (EHR)-linked biobank of over 50,000 participants from the Mount Sinai Health System (MSHS) in New York, NY. T2D cases in BioMe were defined as individuals meeting at least two of the following three criteria: 1) ICD10 code for T2D (code E11 and/or O24.1), 2) a blood value in keeping with diabetes (median HbA1c value greater than or equal to 6.5% and/or median random glucose greater than or equal to 200 mg/dL), and 3) a prescription for any diabetic medication. Individuals were excluded from the case pool if they had an ICD10 code for T1D (code E10 and/or O24.0) or if they had a record of having received an outpatient prescription for insulin (and no record of other antidiabetic medication). Controls were defined as individuals with no ICD10 codes for any type of diabetes mellitus or a family history of diabetes, median HbA1c value of less than 5.7%, median random glucose of less than 200 mg/dL, no oral glucose tolerance test in pregnancy exceeding a diagnostic threshold for gestational diabetes, and no record of a prescription for any diabetic medication.

The Malmö Diet and Cancer Study (MDCS) is a prospective study of ∼53,000 adults living in Malmö, Sweden^3^. T2D cases in MDCS were defined as individuals meeting at least two of the following four criteria: 1) ICD10 code for T2D (code E11 and/or O24.1) or T2D noted in diabetes registries, 2) a blood value in keeping with diabetes (HbA1c value greater than or equal to 6.5% and/or fasting glucose greater than or equal to 126 mg/dL), 3) a prescription for non-insulin diabetic medication, and 4) a record of a non-specific diabetes event (e.g. reported at baseline, or extracted from a registry) with an age at diagnosis, or start of treatment, of greater than or equal to 35 years. Individuals were excluded from the case pool if they had an ICD10 code for T1D (code E10 and/or O24.0), or T1D noted in a diabetes registry, or if they had a record of having received insulin with no record of other antidiabetic medication. Controls were defined as individuals with no ICD10 codes for any type of diabetes mellitus, no family history of diabetes, no other variables indicating a potential diagnosis of diabetes, HbA1c value of less than 5.7%, fasting glucose of less than 100 mg/dL, and no record of a prescription for any diabetic medication.

### Association analyses

Rank-based inverse normal transformed (RINT) quantitative measures (including all subjects and sex-stratified models) with non-missing phenotype information were assessed using an additive mixed model implemented in REGENIE v2^4^. Prior to normalization, traits were adjusted for a standard set of covariates including age, age^2^, sex, age×sex, age^2^×sex, 10 common variant genetic principal components and 20 genetic principal components derived from rare variants. Binary outcomes were similarly adjusted for age, age^2^, sex, age×sex, age^2^×sex, 10 common variant genetic principal components and 20 genetic principal components derived from rare variants and tested for association using a generalized mixed model implemented in REGENIE v2. Following analysis within each cohort, we performed inverse variance-weighted meta-analysis for T2D using METAL.

### GHS serum Call Back Study

As an orthogonal biochemical assessment of the EHS reported blood analyte data, a serum callback study was designed to evaluate serum zinc, blood glucose, insulin synthesis (proinsulin/insulin ratio) and clearance (insulin/c-peptide ratio) in heterozygous carriers of SLC39A5 pLOF variants. Carriers of pLOF variants in SLC39A5 among exome-sequenced participants of European ancestry in the Regeneron Genetics Center-Geisinger Health System DiscovEHR study were included. Controls included non-carriers of pLOF variants in SLC39A5 and SLC30A8 or the common T2D risk variant rs13266634 in SLC30A8 and non-carrier first degree relatives of study subjects. Participants with T1D or T2D diagnoses were excluded. Furthermore, two non-carriers were selected for each carrier matching sex, age (+/- 5years) and BMI (+/- 5). A total of 22 SLC39A5 LOF variants in 131 carriers and 262 matched non-carriers were identified, however sample (frozen fasting serum) availability limited analyses to ∼250 non-carriers and ∼90 carriers as shown in Supplementary Table 1. Serum insulin was measured using Human Insulin ELISA kit (Millipore, EZHI-14BK), proinsulin using Human Total Proinsulin ELISA kit (Millipore, EZHPI-15BK), and c-peptide using Human c-peptide ELISA kit (Abcam, ab178611). Serum zinc was measures using flame atomic absorption spectroscopy as described below. Blood glucose was evaluated using ADVIA Chemistry Glucose Hexokinase_3 reagents (REF 050011429) on a Siemens ADVIA Chemistry XPT analyzer.

### Generation of *Slc39a5* loss of function mice

The genetically engineered *Slc39a5*^-/-^ mouse strain was created using Regeneron’s VelociGene^®^ technology^5, 6^. Briefly, C57Bl/6NTac embryonic stem cells (ESC) were targeted for ablation of a portion of *Slc39a5*, beginning just after the initiating ATG and ending 5 base pairs before the 3’ end of coding exon 2. This region contains the SLC39A5 signal peptide and much of the N-terminal extracellular domain. A lacZ reporter module was inserted in frame with *Slc39a5*’s initiating Methionine codon, followed by a self-deleting fLoxed neomycin resistance (neo) cassette for selection in mouse C57BL/6NTac embryonic stem cells. The targeted cells were microinjected into 8-cell embryos from Charles River Laboratories Swiss Webster albino mice, yielding F0 VelociMice^®^ that were 100% derived from the targeted cells^5^. These mice were subsequently bred to F1, at which point the self deleting neo cassette was also removed in the male germline. F1 heterozygotes were utilized to generate experimental cohorts, including Slc39a5^-/+^ heterozygous mice and wild-type littermates that were used as controls; this line was maintained in Regeneron’s animal facility in the C57Bl/6NTac genetic background throughout the study.

### Animal studies

Mice homozygous for *Slc39a5* loss of function and wild-type littermates were co-housed in a controlled environment (12hr light/dark cycle, 22 ± 1°C, 60-70% humidity) and fed ad-libitum. All studies were performed in both sexes. For HFFD study, ten-week-old mice were fed HFFD diet (46kcal% Fat, 30kcal% Fructose, TestDiet 5WK9) or control diet (TestDiet 58Y2) for 30 weeks. For NASH study, ten-week-old mice were fed NASH diet (40kcal% Fat, 20 kcal% Fructose and 2% Cholesterol, ResearchDiets D09100310) or control diet (ResearchDiets D09100304) for 40 weeks. Both HFFD and NASH diets contain ∼34ppm zinc as described in diet spec sheets, and further confirmed by flame atomic absorption spectrometry. *Slc39a5*^-/-^;*Lepr*^-/-^ mice and corresponding control mice (*Slc39a5*^+/+^;*Lepr*^-/-^, *Slc39a5*^-/-^;*Lepr*^+/+^, and *Slc39a5*^+/+^;*Lepr*^+/+^ mice) were fed a normal chow (LabDiet 5053, containing 87ppm zinc) for 34 weeks. All mice used in this study were housed in pathogen-free environment at Regeneron Pharmaceuticals Inc. animal research facility. Sterile water and show were given ad libitum.

### Ethics Statement

All experimental protocols in mice including anesthesia and tissue sampling procedures performed in this study, were approved by Regeneron Pharmaceuticals Inc. Institutional Animal Care and Use Committee (IACUC) under protocol number 430 and the US Animal Welfare Act.

### Serum Analysis

Sera were collected upon an overnight fast (16 hours). The liver and lipid profile were analyzed using Siemens ADVIA Chemistry XPT analyzer which is maintained and operated according to Siemens’ guidelines. The liver and lipid profile contains the following reagents: Alanine Aminotransferase (ALT, Siemens REF 03036926), Aspartate Aminotransferase (AST, Siemens REF 07499718), Cholesterol (CHOL, Siemens REF 10376501), Direct HDL Cholesterol (DHDL, Siemens REF 07511947), LDL Cholesterol Direct (DLDL, Siemens REF 09793248), Non-Esterified Fatty Acids (NEFA, Wako 999-34691, 995-34791, 991-34891, 993-35191), Triglycerides (TRIG, Siemens REF 10335892. When mixed with sample, reagents undergo a colorimetric change proportional to the concentration of the specific analyte. The absorbance is then measured with a halogen light source and used to determine concentration. Serum was also collected for ELISA analysis of proinsulin (Mercodia, #10-1232-01) per manufacturer’s guidelines. Briefly, samples were incubated with enzyme conjugate at room temperature for two hours and washed. Substrate TMB was added and the reaction was allowed to proceed for 30 minutes at room temperature before stop solution was applied. Optical density was read at 450 nm. Luminex Metabolic panel serum analyses of insulin, c-peptide were performed using a Mouse Metabolic Hormone Magnetic Bead Panel (Millipore, MMHMAG-44K). Experimental protocols for the sample collection, storage, preparation of reagents for immunoassay and immunoassay procedure, followed the specific instructions of the MMHMAG-44K mouse panel supplier. Results were read on a Luminex 200 analyzer with XX software (Xponent/Analyst version 4.2) used for data analysis. Insulin profile was also analyzed in serum collected from mice at fed state.

### Metal ion quantification

Assays were performed by the Louisiana Animal Disease Diagnostics Laboratory with an Agilent Technologies 240 FS Atomic Absorption Spectrometer, in flame mode. Serum samples are quantitatively diluted in deionized water and subsequently analyzed. For the serum samples a Seronorm Trace Elements Serum (L-2) is used as reference. First tissue samples are weighed and digested in nitric acid overnight at 85°C. The following day, the samples are cooled down to room temperature and quantitatively transferred to polystyrene tubes with deionized water, and subsequently analyzed. For all tissue samples, a bovine liver standard reference material (SRM 1577c) from the National Institute of Standards and Technology was used as reference.

### Liver histology and histopathologic analysis

Explanted liver samples were fixed in 10% phosphate buffered formalin acetate at 4⁰C overnight, thoroughly rinsed in phosphate-buffered saline and transferred to 70% ethanol. Histology was performed by HistoWiz Inc. and Histoserv Inc. using standard operating procedures and fully automated workflow. Samples were embedded in paraffin wax and sectioned (5 µm). Prior to staining, slides were deparaffinized in xylene and hydrated with graded alcohols and finally water. Slides were then stained with either hematoxylin & eosin (H&E) or Picrosirius Red. Immunohistochemistry was performed on a Bond Rx autostainer (Leica Biosystems) with heat-induced epitope retrieval. Slides were incubated with primary antibodies F4/80 (Thermo, #14-4801-82), α-smooth muscle actin (Abcam, #ab5694) and Bond Polymer Refine Detection (Leica Biosystems) was used per manufacturer’s protocol. Following staining, slides were dehydrated and coverslipped using a TissueTek-Prisma and Coverslipper (Sakura). Whole slide scanning (40x) was performed on an Aperio AT2 (Leica Biosystems). For lipid staining, samples were frozen in O.C.T. compound (Tissue-Tek, #4583) and 5µm thick sections were used. Slides were stained with Oil Red O (ORO) and Mayers hematoxylin and mounted with glycerin jelly. NAFLD scoring was performed by one external pathologist (provided by Histowiz) and one internal pathologist blinded to the samples, according to criteria described by Liang et al^7^. Macrovesicular steatosis (H&E, ORO), hepatocyte hypertrophy (H&E), inflammation (H&E, F4/80) and fibrosis (PSR) were scored ranging from 0 to 3. NAFLD activity score is the sum of steatosis, hepatocyte hypertrophy and inflammation scores. Steatosis-activity-fibrosis score is the sum of NAFLD activity score and fibrosis score.

### Hepatic Triglyceride assay

Lipids were extracted from liver samples using Folch method^8^ and solubilized as described earlier^9^. The levels of triglyceride were measured using Pointe triglyceride (GPO) reagent set (MedTest Dx, #T7532) and normalized to wet tissue weight.

### Glucose and insulin tolerance tests

An oral glucose tolerance test was administered upon an overnight fast (16 hours) with free access to water. Dextrose (Hospira Inc, NDC 0409-4902-34) was administered by oral gavage per 2g/kg of body weight. Blood glucose was evaluated at defined time points (0, 15, 30, 60 and 120 minutes) using AlphaTrak blood glucose monitoring system (Zoetis United States, Parsippany NJ) by sampling blood from the lateral tail vein. Insulin tolerance tests were performed after a 4 hour fast by administering 1.0U/kg of body weight of Humulin R (Eli Lilly, #HI-213) by intra-peritoneal injection. Blood glucose was again evaluated at defined timepoints with the AlphaTrak blood glucose monitoring system by sampling blood from the lateral tail vein.

### HOMA-IR

Homeostatic model assessment of insulin resistance (HOMA-IR) indicates level of insulin sensitivity by taking into account the relationship between glucose and insulin. HOMA-IR was calculated according to the formula: fasting insulin (microU/L) x fasting glucose (nmol/L)/22.5.

### Immunoblotting

For biochemical analysis, liver samples were harvested and immediately snap frozen in liquid nitrogen. Protein was later extracted using RIPA buffer (Cell signaling technology, #9806) with Halt Protease & Phosphatase Inhibitor Cocktail (ThermoFisher Scientific, #78440). Protein concentration was determined using Pierce TM BCA protein assay kit (Thermo Scientific, #23225). Five micrograms of protein of each sample were separated in NuPAGE 4-12% Bis-Tris protein gel (Invitrogen, #WG1403BOX), and transferred to nitrocellulose membrane using Trans-Blot® Turbo™ Transfer System (BioRad). The membranes were blocked with 5% non-fat dry milk (BioRad, #9999) for 1 hour at room temperature before incubated with primary antibody overnight at 4°C. Antibodies were purchased from Cell Signaling Technology, phospho-AKT (Ser473, #4060), AKT (#9272), phospho-AMPKα (Thr172, #2535), AMPKα (#5831), phospho-LKB1 (Ser428, #3482), LKB1 (#3047), phospho-ACC (Ser79, #3661), ACC (#3676), FASN (#3189), HRP-linked anti-rabbit IgG (#7074) and HRP-linked anti-mouse IgG (#7076). Antibodies for G6PC (Invitrogen, #PA5-42541), SLC39A5 and β-actin (Sigma #5441) were used. For detection of SLC39A5 protein, liver samples were immunoprecipitated using Pierce Protein A/G Magnetic Beads and anti-SLC39A5 antibodies (Invitrogen, #88803, 42522), and eluted for western blot analysis. All membranes were washed before incubation with HRP-linked secondary antibody for one hour at room temperature. Blots were developed using SuperSignal West Femto Substrate (ThermoFisher Scientific, #34095). Signals were captured using G:Box Mini 9 (Syngene). Densitometry analysis of immunoblots were performed using ImageJ.

### Gene expression analysis

Tissues were preserved in RNAlater solution immediately following harvest. Total RNA was purified using MagMAX™-96 for Microarrays Total RNA Isolation Kit (Invitrogen, #AM1839) according to manufacturer’s specifications. Genomic DNA was removed using MagMAX™Turbo™DNase Buffer and TURBO DNase from the MagMAX kit listed above. mRNA (Up to 2.5ug) was reverse-transcribed into cDNA using SuperScript® VILO™ Master Mix (Invitrogen, #11755500). cDNA was diluted to 0.5-5ng/uL. 2.5-25ng cDNA input was amplified with the SensiFAST Hi-ROX MasterMix (BIOLINE, #CSA-01113) using the ABI 7900HT Sequence Detection System (Applied Biosystems). The sequences of primers are as follows: *Slc39a5* (F 5’-CGAGCCTAGACCTCTTCCA-3’, R 5’-GGGAGCCATTCAGACAATCC-3’), *Mt1* (F 5’-CAAGTGCACCTCCTGCAAGAAG-3’, R 5’-CACAGCCCTGGGCACATTT-3’), *Mt2* (F 5’-GACCCCAACTGCTCCTGTG-3’, R 5-‘CTTGCAGGAAGTACATTTGCATTG-3’), *G6pc* (F5’-GGTCGTGGCTGGAGTCTTG-3’, R 5’-CCGGAGGCTGGCATTGTAG-3’) and *Fasn* (Thermofisher Scientific #Mm00662319_m1).

### Human primary hepatocyte culture

Human Plateable Hepatocytes were purchased from Invitrogen (#HMCPP5, Lot No HPP1881027 and HPP1878738) and used according to manufacturer protocol. These Hepatocytes are a pooled population of primary hepatocytes produced by combining cells from 5 individual donors. All reagents and materials were purchased from Invitrogen. Briefly, cryopreserved hepatocytes were thawed in hepatocyte thawing medium (#CM7500). Hepatocytes were centrifuged and resuspend in plating medium, Williams’ Medium E (#A1217601) with hepatocyte plating supplement (#CM3000). Hepatocytes were directly plated in collagen I coated 24-well plate (#A1142802). After 6 hours incubation, media were replaced with incubation medium, Williams’ Medium E with hepatocyte maintenance supplement (#CM4000). Next day, hepatocytes were treated with ZnCl_2_ or MgCl_2_ at the concentrations of 100, 200 and 400uM in incubation medium for four hours. Magnesium was used as a negative control, given that zinc and magnesium have opposite roles in the activation of protein tyrosine phosphatase 1B ^10^. In addition, Okadaic acid (OA) and Metformin (Met) were used as positive controls. OA is an inhibitor of the serine/threonine protein phosphatases (PP2A and PP1), resulting in an elevation of p.AMPKα Thr172 and p.AKT Ser473 levels in hepatocytes ^11, 12^. Metformin is an antidiabetic drug that induces phosphorylation of AMPK in liver ^13^. Protein lysates were collected using RIPA buffer with Halt Protease & Phosphatase Inhibitor and subjected for immunoblotting. Cell Viability assay was performed using CellTiter 96® AQueous One Solution Cell Proliferation Assay (MTS) assay (Promega, #G3580) per manufacturer’s protocol.

### Generation of Slc39a5 plasmids

Mouse *Slc39a5* ORF sequence was cloned into pIRES2 DsRed-Express2 vector (Clontech, #PT4079-5). Constructs of SLC39A5 variants were generated using site-directed mutagenesis method, with oligos for Y47X (F 5’CCCATTCTCGCCCTACAGGCCAAACAGCTG-3’, R 5-CAGCTGTTTGGCCTGTAGGGCGAGAATGGG-3’), R311X (F 5’-GGCCTGAGCCCTCAGTGCCGCAAAAGC-3’, R 5’-GCTTTTGCGGCACTGAGGGCTCAGGCC-3’), R322X (F 5’-GTTTCGAGATTCCTTCATTTTCGCCTGCAGCATCT-3’, R 5’-AGATGCTGCAGGCGAAAATGAAGGAATCTCGAAAC-3’) and M304T (F 5’-AAAGCCCCAGCGTGTTCTCCAGCACAAAGAGCA-3’, R 5’-TGCTCTTTGTGCTGGAGAACACGCTGGGGCTTT-3’). Mutagenesis was confirmed by Sanger sequencing.

### Membrane localization of SLC39A5 using flow cytometry

HEK293 cells were plated on a 10cm dish at a density of 25,000 cells/cm^2^ and incubated at 37°C at 5% CO_2_ overnight in high glucose DMEM with 10% FBS and 1% Penn Strep (Gibco, #11965092). Cells were transfected the next day with 10ug plasmid DNA using Xtremegene HP (Roche, #06366244001) in Opti-MEM (Gibco, #31985-070). Transfection complexes were incubated at room temperature for 20 minutes and added dropwise to the cells. Cells were incubated at 37°C. A single media change was performed after 48 hours using Gibco DMEM high glucose and no other supplements. Following 24 hours of incubation at 37°C cells were washed with DPBS and dissociated with Cell dissociation Buffer (Gibco, #13151-014). Cells were resuspended in 0.5% BSA (Sigma, #A7030) in DPBS and centrifuged at 200g for 5 minutes. BD Biosciecnes CytoFix Fixation buffer (BD, #554655) was added to each sample and incubated at 4C for 20 minutes. Cells were washed and stained with either anti-human SLC39A5 (Sigma, #SAB1408465) or isotype control followed by secondary Alexa 488 (ThermoFisher, #A28175). Cells were washed and re-suspended in 0.5% BSA in DPBS. All samples were analyzed on a BD FACS CantoII.

### *MRE*-luc assay

HEK293 cells were plated in 96-well plate at a density of 22,000 cells/well. Cells were transfected with MRE-luc (Promega pGL4.40), hRluc (Promega pGL4.75) and *Slc39a5* constructs with XtremeGene HP (Roche) transfection reagent. MRE-binding transcription factor 1 (MTF1) is responsible for expression of metallothioneins (MTs) in response to zinc. After 24 hours, medium was changed with 0.5% charcoal stripped FBS in DMEM + Glutamax (Life Technologies, #10569010). Twenty-four hours later, the cultures were exposed to Zn^2+^ for 6 hours. Luciferase activity was measured using Dual-Glo luciferase assay system (Promega, #E2920). Results were expressed as the relation of firefly luciferase activity to Renilla luciferase activity.

### Live staining & Immunofluorescence

HEK293 cells were plated on Ibidi chamber slides (Ibidi, #80427) coated with 100µg/mL of poly-L-lysine. Live cells staining was performed by diluting primary antibody (Polyclonal anti-SLC39A5, Sigma SAB1408465, 1:100) in cold culture medium and incubated with the cells for 2 hours at 4°C. Slides were washed with cold PBS 3x 10 minutes at 4°C with gentle shaking. Cells were fixed with 2% paraformaldehyde at room temperature for 5 minutes and washed with PBS. Wheat germ agglutinin in HBSS was added to cells for 10 minutes at room temperature and then blocked with 5% goat serum. Secondary antibody was applied for 45 minutes at room temperature. Cells were washed and mounted with ProLong diamond antifade solution and imaged using Zeiss AxioObserver LSM880.

### Hepatic β-hydroxybutyrate assay

β-OHB (Sigma, #MAK041) was measured in mouse liver tissue by colorimetric assays per manufacturer’s guidelines. Briefly, liver samples (10mg/sample) were homogenized in 4 volumes of cold β-OHB assay buffer. Samples were centrifuged at 13,000g for 10 minutes at 4°C and supernatant was used for analyses. Concentration is determined by a coupled enzyme reaction which results in a colorimetric (450nm) product proportional to the β-OHB present.

### Protein Phosphatase Assay

Cultured hepatocyte lysates and tissue homogenates were prepared by Lysis Reagent (Invitrogen, #M10510) with protease inhibitor cocktail (Sigma, #P8340). Protein lysates were centrifuged at 4⁰C for 15 minutes and protein concentration was determined using Pierce TM BCA protein assay kit. Phosphatase activity was measured using RediPlate™ 96 EnzChek™ Tyrosine Phosphatase and Serine/Threonine Phosphatase Assay Kit (Invitrogen, #R22067, R33700) per manufacturer’s instructions. Samples were incubated in RediPlate for 60 mins before reading by fluorescence microplate with excitation/emission at 358/455nm (Spectramax M4, Molecular Devices).

### Superoxide Dismutase (SOD) assay

SOD activity was measured in mouse liver tissues using SOD activity kit (ENZO, #ADI-900-157). Briefly, liver samples (30mg/sample) were homogenized in 10 volumes of cell extraction buffer. Samples were centrifuged at 10,000g for 10 minutes at 4°C and supernatant was used for analysis. Protein concentration was determined by Pierce TM BCA protein assay kit (Thermo Scientific, #23225). Samples were diluted to 50ug/25ul for assay, according to protocol.

### Statistical analysis

Results are shown as box plots with individual values. Statistical analysis was performed using GraphPad Prism 8 software. Analysis for *Slc39a5^-/-^*; *Lepr^-/-^* mice and corresponding control mice was performed using one-way ANOVA, followed by post hoc Tukey’s tests. Analysis for HFFD and NASH mouse studies was performed using two-way ANOVA, followed by post hoc Tukey’s tests. Statistical significance reported when p<0.05. Sample sizes, statistical test and significance are described in each figure legend.

## References

1. M. J. Jackson, D. A. Jones, R. H. Edwards, Tissue zinc levels as an index of body zinc status. Clin Physiol 2, 333–343 (1982).

2. R. E. Dempski, The cation selectivity of the ZIP transporters. Curr Top Membr 69, 221–245 (2012).

3. P. Ranasinghe, W. S. Wathurapatha, P. Galappatthy, P. Katulanda, R. Jayawardena, G. R. Constantine, Zinc supplementation in prediabetes: A randomized double-blind placebo-controlled clinical trial. J Diabetes 10, 386–397 (2018).

4. J. Flannick, G. Thorleifsson, N. L. Beer, S. B. Jacobs, N. Grarup, N. P. Burtt, A. Mahajan, C. Fuchsberger, G. Atzmon, R. Benediktsson, J. Blangero, D. W. Bowden, I. Brandslund, J. Brosnan, F. Burslem, J. Chambers, Y. S. Cho, C. Christensen, D. A. Douglas, R. Duggirala, Z. Dymek, Y. Farjoun, T. Fennell, P. Fontanillas, T. Forsen, S. Gabriel, B. Glaser, D. F. Gudbjartsson, C. Hanis, T. Hansen, A. B. Hreidarsson, K. Hveem, E. Ingelsson, B. Isomaa, S. Johansson, T. Jorgensen, M. E. Jorgensen, S. Kathiresan, A. Kong, J. Kooner, J. Kravic, M. Laakso, J. Y. Lee, L. Lind, C. M. Lindgren, A. Linneberg, G. Masson, T. Meitinger, K. L. Mohlke, A. Molven, A. P. Morris, S. Potluri, R. Rauramaa, R. Ribel-Madsen, A. M. Richard, T. Rolph, V. Salomaa, A. V. Segre, H. Skarstrand, V. Steinthorsdottir, H. M. Stringham, P. Sulem, E. S. Tai, Y. Y. Teo, T. Teslovich, U. Thorsteinsdottir, J. K. Trimmer, T. Tuomi, J. Tuomilehto, F. Vaziri-Sani, B. F. Voight, J. G. Wilson, M. Boehnke, M. I. McCarthy, P. R. Njolstad, O. Pedersen, T. D. C. Go, T. D. G. Consortium, L. Groop, D. R. Cox, K. Stefansson, D. Altshuler, Loss-of-function mutations in SLC30A8 protect against type 2 diabetes. Nat Genet 46, 357–363 (2014).

5. O. P. Dwivedi, M. Lehtovirta, B. Hastoy, V. Chandra, N. A. J. Krentz, S. Kleiner, D. Jain, A. M. Richard, F. Abaitua, N. L. Beer, A. Grotz, R. B. Prasad, O. Hansson, E. Ahlqvist, U. Krus, I. Artner, A. Suoranta, D. Gomez, A. Baras, B. Champon, A. J. Payne, D. Moralli, S. K. Thomsen, P. Kramer, I. Spiliotis, R. Ramracheya, P. Chabosseau, A. Theodoulou, R. Cheung, M. van de Bunt, J. Flannick, M. Trombetta, E. Bonora, C. B. Wolheim, L. Sarelin, R. C. Bonadonna, P. Rorsman, B. Davies, J. Brosnan, M. I. McCarthy, T. Otonkoski, J. O. Lagerstedt, G. A. Rutter, J. Gromada, A. L. Gloyn, T. Tuomi, L. Groop, Loss of ZnT8 function protects against diabetes by enhanced insulin secretion. Nat Genet 51, 1596–1606 (2019).

6. M. Baron, A. Veres, S. L. Wolock, A. L. Faust, R. Gaujoux, A. Vetere, J. H. Ryu, B. K. Wagner, S. S. Shen-Orr, A. M. Klein, D. A. Melton, I. Yanai, A Single-Cell Transcriptomic Map of the Human and Mouse Pancreas Reveals Inter- and Intra-cell Population Structure. Cell Syst 3, 346–360 e344 (2016).

7. M. J. Muraro, G. Dharmadhikari, D. Grun, N. Groen, T. Dielen, E. Jansen, L. van Gurp, M. A. Engelse, F. Carlotti, E. J. de Koning, A. van Oudenaarden, A Single-Cell Transcriptome Atlas of the Human Pancreas. Cell Syst 3, 385–394 e383 (2016).

8. F. Wang, B. E. Kim, M. J. Petris, D. J. Eide, The mammalian Zip5 protein is a zinc transporter that localizes to the basolateral surface of polarized cells. J Biol Chem 279, 51433–51441 (2004).

9. J. Geiser, R. C. De Lisle, G. K. Andrews, The zinc transporter Zip5 (Slc39a5) regulates intestinal zinc excretion and protects the pancreas against zinc toxicity. PLoS One 8, e82149 (2013).

10. X. Wang, H. Gao, W. Wu, E. Xie, Y. Yu, X. He, J. Li, W. Zheng, X. Wang, X. Cao, Z. Meng, L. Chen, J. Min, F. Wang, The zinc transporter Slc39a5 controls glucose sensing and insulin secretion in pancreatic beta-cells via Sirt1- and Pgc-1alpha-mediated regulation of Glut2. Protein Cell 10, 436–449 (2019).

11. J. Dufner-Beattie, Y. M. Kuo, J. Gitschier, G. K. Andrews, The adaptive response to dietary zinc in mice involves the differential cellular localization and zinc regulation of the zinc transporters ZIP4 and ZIP5. J Biol Chem 279, 49082–49090 (2004).

12. B. W. Huang, M. T. Chiang, H. T. Yao, W. Chiang, The effect of high-fat and high-fructose diets on glucose tolerance and plasma lipid and leptin levels in rats. Diabetes Obes Metab 6, 120–126 (2004).

13. A. J. King, The use of animal models in diabetes research. Br J Pharmacol 166, 877–894 (2012).

14. C. Tabula Muris, A single-cell transcriptomic atlas characterizes ageing tissues in the mouse. Nature 583, 590–595 (2020).

15. Y. Xin, J. Kim, M. Ni, Y. Wei, H. Okamoto, J. Lee, C. Adler, K. Cavino, A. J. Murphy, G. D. Yancopoulos, H. C. Lin, J. Gromada, Use of the Fluidigm C1 platform for RNA sequencing of single mouse pancreatic islet cells. Proc Natl Acad Sci U S A 113, 3293–3298 (2016).

16. H. Tilg, A. R. Moschen, M. Roden, NAFLD and diabetes mellitus. Nat Rev Gastroenterol Hepatol 14, 32–42 (2017).

17. D. Garcia, K. Hellberg, A. Chaix, M. Wallace, S. Herzig, M. G. Badur, T. Lin, M. N. Shokhirev, A. F. M. Pinto, D. S. Ross, A. Saghatelian, S. Panda, L. E. Dow, C. M. Metallo, R. J. Shaw, Genetic Liver-Specific AMPK Activation Protects against Diet-Induced Obesity and NAFLD. Cell Rep 26, 192–208 e196 (2019).

18. P. M. Titchenell, W. J. Quinn, M. Lu, Q. Chu, W. Lu, C. Li, H. Chen, B. R. Monks, J. Chen, J. D. Rabinowitz, M. J. Birnbaum, Direct Hepatocyte Insulin Signaling Is Required for Lipogenesis but Is Dispensable for the Suppression of Glucose Production. Cell Metab 23, 1154–1166 (2016).

19. E. Bellomo, K. Birla Singh, A. Massarotti, C. Hogstrand, W. Maret, The metal face of protein tyrosine phosphatase 1B. Coord Chem Rev 327-328, 70–83 (2016).

20. Y. Xiong, X. P. Jing, X. W. Zhou, X. L. Wang, Y. Yang, X. Y. Sun, M. Qiu, F. Y. Cao, Y. M. Lu, R. Liu, J. Z. Wang, Zinc induces protein phosphatase 2A inactivation and tau hyperphosphorylation through Src dependent PP2A (tyrosine 307) phosphorylation. Neurobiol Aging 34, 745–756 (2013).

21. L. Xian, S. Hou, Z. Huang, A. Tang, P. Shi, Q. Wang, A. Song, S. Jiang, Z. Lin, S. Guo, X. Gao, Liver-specific deletion of Ppp2calpha enhances glucose metabolism and insulin sensitivity. Aging (Albany NY) 7, 223–232 (2015).

22. M. Delibegovic, D. Zimmer, C. Kauffman, K. Rak, E. G. Hong, Y. R. Cho, J. K. Kim, B. B. Kahn, B. G. Neel, K. K. Bence, Liver-specific deletion of protein-tyrosine phosphatase 1B (PTP1B) improves metabolic syndrome and attenuates diet-induced endoplasmic reticulum stress. Diabetes 58, 590–599 (2009).

23. S. L. Friedman, B. A. Neuschwander-Tetri, M. Rinella, A. J. Sanyal, Mechanisms of NAFLD development and therapeutic strategies. Nat Med 24, 908–922 (2018).

24. D. N. Marreiro, M. Fisberg, S. M. Cozzolino, Zinc nutritional status in obese children and adolescents. Biol Trace Elem Res 86, 107–122 (2002).

25. M. K. Mohammad, Z. Zhou, M. Cave, A. Barve, C. J. McClain, Zinc and liver disease. Nutr Clin Pract 27, 8–20 (2012).

26. A. Hosui, E. Kimura, S. Abe, T. Tanimoto, K. Onishi, Y. Kusumoto, Y. Sueyoshi, K. Matsumoto, M. Hirao, T. Yamada, N. Hiramatsu, Long-Term Zinc Supplementation Improves Liver Function and Decreases the Risk of Developing Hepatocellular Carcinoma. Nutrients 10, (2018).

27. E. Bellomo, A. Abro, C. Hogstrand, W. Maret, C. Domene, Role of Zinc and Magnesium Ions in the Modulation of Phosphoryl Transfer in Protein Tyrosine Phosphatase 1B. J Am Chem Soc 140, 4446–4454 (2018).

28. S. Lee, G. Chanoit, R. McIntosh, D. A. Zvara, Z. Xu, Molecular mechanism underlying Akt activation in zinc-induced cardioprotection. Am J Physiol Heart Circ Physiol 297, H569–575 (2009).

29. S. Liangpunsakul, M. S. Sozio, E. Shin, Z. Zhao, Y. Xu, R. A. Ross, Y. Zeng, D. W. Crabb, Inhibitory effect of ethanol on AMPK phosphorylation is mediated in part through elevated ceramide levels. Am J Physiol Gastrointest Liver Physiol 298, G1004–1012 (2010).

30. S. Galic, C. Hauser, B. B. Kahn, F. G. Haj, B. G. Neel, N. K. Tonks, T. Tiganis, Coordinated regulation of insulin signaling by the protein tyrosine phosphatases PTP1B and TCPTP. Mol Cell Biol 25, 819–829 (2005).

31. N. Krishnan, K. F. Konidaris, G. Gasser, N. K. Tonks, A potent, selective, and orally bioavailable inhibitor of the protein-tyrosine phosphatase PTP1B improves insulin and leptin signaling in animal models. J Biol Chem 293, 1517–1525 (2018).

32. T. C. Chen, D. I. Benjamin, T. Kuo, R. A. Lee, M. L. Li, D. J. Mar, D. E. Costello, D. K. Nomura, J. C. Wang, The glucocorticoid-Angptl4-ceramide axis induces insulin resistance through PP2A and PKCzeta. Sci Signal 10, (2017).

33. M. Y. Wang, R. H. Unger, Role of PP2C in cardiac lipid accumulation in obese rodents and its prevention by troglitazone. Am J Physiol Endocrinol Metab 288, E216–221 (2005).

34. A. Pal, T. M. Barber, M. Van de Bunt, S. A. Rudge, Q. Zhang, K. L. Lachlan, N. S. Cooper, H. Linden, J. C. Levy, M. J. Wakelam, L. Walker, F. Karpe, A. L. Gloyn, PTEN mutations as a cause of constitutive insulin sensitivity and obesity. N Engl J Med 367, 1002–1011 (2012).

35. R. C. Tsou, K. K. Bence, The Genetics of PTPN1 and Obesity: Insights from Mouse Models of Tissue-Specific PTP1B Deficiency. J Obes 2012, 926857 (2012).

36. M. Regnier, A. Polizzi, S. Smati, C. Lukowicz, A. Fougerat, Y. Lippi, E. Fouche, F. Lasserre, C. Naylies, C. Betoulieres, V. Barquissau, E. Mouisel, J. Bertrand-Michel, A. Batut, T. A. Saati, C. Canlet, M. Tremblay-Franco, S. Ellero-Simatos, D. Langin, C. Postic, W. Wahli, N. Loiseau, H. Guillou, A. Montagner, Hepatocyte-specific deletion of Pparalpha promotes NAFLD in the context of obesity. Sci Rep 10, 6489 (2020).

37. J. J. Zhang, J. J. Hao, Y. R. Zhang, Y. L. Wang, M. Y. Li, H. L. Miao, X. J. Zou, B. Liang, Zinc mediates the SREBP-SCD axis to regulate lipid metabolism in Caenorhabditis elegans. J Lipid Res 58, 1845–1854 (2017).

38. Y. Bai, J. G. McCoy, E. J. Levin, P. Sobrado, K. R. Rajashankar, B. G. Fox, M. Zhou, X-ray structure of a mammalian stearoyl-CoA desaturase. Nature 524, 252–256 (2015).

39. D. M. Valenzuela, A. J. Murphy, D. Frendewey, N. W. Gale, A. N. Economides, W. Auerbach, W. T. Poueymirou, N. C. Adams, J. Rojas, J. Yasenchak, R. Chernomorsky, M. Boucher, A. L. Elsasser, L. Esau, J. Zheng, J. A. Griffiths, X. Wang, H. Su, Y. Xue, M. G. Dominguez, I. Noguera, R. Torres, L. E. Macdonald, A. F. Stewart, T. M. DeChiara, G. D. Yancopoulos, High-throughput engineering of the mouse genome coupled with high-resolution expression analysis. Nat Biotechnol 21, 652–659 (2003).

## References

1. Eastwood SV, Mathur R, Atkinson M, Brophy S, Sudlow C, Flaig R, et al. Algorithms for the Capture and Adjudication of Prevalent and Incident Diabetes in UK Biobank. PLoS One 2016;11:e0162388.

2. Lotta LA, Wittemans LBL, Zuber V, Stewart ID, Sharp SJ, Luan J, et al. Association of Genetic Variants Related to Gluteofemoral vs Abdominal Fat Distribution With Type 2 Diabetes, Coronary Disease, and Cardiovascular Risk Factors. JAMA 2018;320:2553–2563.

3. Berglund G, Elmstahl S, Janzon L, Larsson SA. The Malmo Diet and Cancer Study. Design and feasibility. J Intern Med 1993;233:45–51.

4. Mbatchou J, Barnard L, Backman J, Marcketta A, Kosmicki JA, Ziyatdinov A, et al. Computationally efficient whole-genome regression for quantitative and binary traits. Nat Genet 2021.

5. Poueymirou WT, Auerbach W, Frendewey D, Hickey JF, Escaravage JM, Esau L, et al. F0 generation mice fully derived from gene-targeted embryonic stem cells allowing immediate phenotypic analyses. Nat Biotechnol 2007;25:91–99.

6. Valenzuela DM, Murphy AJ, Frendewey D, Gale NW, Economides AN, Auerbach W, et al. High-throughput engineering of the mouse genome coupled with high-resolution expression analysis. Nat Biotechnol 2003;21:652–659.

7. Liang W, Menke AL, Driessen A, Koek GH, Lindeman JH, Stoop R, et al. Establishment of a general NAFLD scoring system for rodent models and comparison to human liver pathology. PLoS One 2014;9:e115922.

8. Folch J, Lees M, Sloane Stanley GH. A simple method for the isolation and purification of total lipides from animal tissues. J Biol Chem 1957;226:497–509.

9. Carr TP, Andresen CJ, Rudel LL. Enzymatic determination of triglyceride, free cholesterol, and total cholesterol in tissue lipid extracts. Clin Biochem 1993;26:39–42.

10. Bellomo E, Abro A, Hogstrand C, Maret W, Domene C. Role of Zinc and Magnesium Ions in the Modulation of Phosphoryl Transfer in Protein Tyrosine Phosphatase 1B. J Am Chem Soc 2018;140:4446–4454.

11. Samari HR, Moller MT, Holden L, Asmyhr T, Seglen PO. Stimulation of hepatocytic AMP-activated protein kinase by okadaic acid and other autophagy-suppressive toxins. Biochem J 2005;386:237–244.

12. Galbo T, Olsen GS, Quistorff B, Nishimura E. Free fatty acid-induced PP2A hyperactivity selectively impairs hepatic insulin action on glucose metabolism. PLoS One 2011;6:e27424.

13. Howell JJ, Hellberg K, Turner M, Talbott G, Kolar MJ, Ross DS, et al. Metformin Inhibits Hepatic mTORC1 Signaling via Dose-Dependent Mechanisms Involving AMPK and the TSC Complex. Cell Metab 2017;25:463–471.

